# Rapid and long-lasting remodelling of the blood transcriptome following bariatric surgery

**DOI:** 10.1101/2025.06.20.25329990

**Authors:** Julien Roux, Denis Seyres, Jessica L Buxton, Dale Handley, Giulia Hofer, Suzanne Alsters, Karl-Heinz Herzig, Marjo-Riitta Jarvelin, Alexandra I Blakemore, Claudia Cavelti-Weder

## Abstract

Bariatric (or metabolic) surgery is the most effective treatment for severe obesity, resulting in sustained weight loss and rapid improvement in metabolic outcomes. However, it is currently unknown what molecular changes are induced by the surgery, how they relate to the health improvements, how early they occur, and whether they are maintained over time.

In this study, we characterized early gene expression changes following surgery in blood samples from the “Personalised Medicine for Morbid Obesity” cohort. We observed widespread changes in gene expression only a few days after surgery. Pathways related to immune response were affected, in particular with a decreased expression of the NF-κB pathway and neutrophil cytotoxic activity genes, suggesting a reduction of the inflammatory state typically associated with obesity. Metabolic signaling was also affected, with decreased expression of genes from pathways related to glucagon and insulin secretion, and key regulators such as the appetite-controlling hormone ghrelin. Comparisons to publicly available transcriptomics datasets showed that beyond these specific changes, bariatric surgery induces a transcriptome-wide reversal of expression changes associated with obesity and with forms of type 2 diabetes related to obesity. Comparisons to transcriptomics studies with longer-term follow-ups after bariatric surgery showed that a large fraction of early-induced expression changes likely persist for several months.

Taken together, we show that previously unreported early changes in blood gene expression after bariatric surgery provide in-depth insights into the resolution of the chronic inflammation associated with severe obesity and its connection with metabolic improvement.

## Introduction

Obesity is a major public health issue with significant societal consequences. It is associated with a chronic state of inflammation, which is thought to lead to the development of comorbidities such as type 2 diabetes mellitus (T2D)[1, 2]. The low-grade inflammation accompanying the progression to T2D is apparent from altered homeostasis of innate and adaptive immune cell types in multiple tissues. An analysis of blood lymphocytes from a cohort of patients with T2D and concomitant obesity showed that the cytotoxic function of CD8+ T cells was impaired compared to matched controls [3]. CD4+ regulatory and helper T cell subsets are also polarized, with depletion of pro-inflammatory Th1 and Th17 subsets at the expense of anti-inflammatory Th2 and Treg subsets [4–9]. At the molecular level, these changes are associated with the activation of the pro-inflammatory NF-κB pathway, enhancing the production of pro-inflammatory cytokines such as TNF, IL-6 and IL-1β, which in turn trigger a cascade of downstream events on immune and non-immune cell types [1, 2, 10].

Metabolic and bariatric surgery (MBS) through “Roux-en-Y’’ gastric bypass (RYGB) or sleeve gastrectomy (SG) is the most effective intervention to treat severe obesity (BMI >40 kg/m^2^, or BMI >35 kg/m^2^ with significant comorbidities)[11]. MBS usually results in substantial and sustained weight loss, and durable improvement in the patient’s metabolic status [12–14], helping patients with concomitant obesity and T2D to achieve long-term remission. For example, a remission rate of 51% was observed at least 5 years after RYGB in a US-based study on 686 patients with T2D [15]. It was even above 70% after 5 years in a study on 635 Danish patients with T2D [16] – but note that the criteria used for the definition of remission differ slightly across studies.

At the immune level, MBS was shown to be associated with a reversal of the inflammatory state associated with obesity and T2D, specifically a rebalancing of the blood Th1/Th2 ratio and a decrease in circulating inflammatory cytokines [17]. This effect is observed rapidly after MBS, leading to the hypothesis that the resolution of inflammation could potentially play a causal role in the improvement of glucose metabolism [12]. However, the available evidence fort such a causal link is conflicting: A meta-analysis of 116 studies demonstrated that levels of serum inflammatory factors can indeed be reduced as early as 1 month after MBS, but there was significant heterogeneity across studies, and few studies included such early follow-up timepoints [18]. Another study on a cohort of 32 patients described a time lag between the metabolic improvement, observed already 3 months after MBS, and the significant decrease of inflammatory factors in plasma, observed only after 6 months [19]. Last, the metabolic improvement after MBS can be strikingly rapid: normoglycaemia (without medication) is often observed in patients with T2D a few days after MBS, by the time of hospital discharge [20, 21], an effect that may be too rapid to be primarily driven by the resolution of inflammation. Overall, few studies have investigated the temporal aspect of the metabolic and immune changes induced by MBS. In particular, little is known about the early effects of the surgery.

This led us to investigate the early molecular changes in blood after MBS. Our study was conducted in the context of a registered clinical trial intended to investigate the mechanisms underlying T2D remission after MBS. We used RNA-sequencing to characterize the transcriptome of matched whole blood samples collected from 24 study participants during pre-surgery assessment and a few days after surgery. To our knowledge, this is the first large-scale investigation of the early molecular changes in blood after MBS.

## Results

### Phenotypic characterisation of the cohort

The present cohort includes a subset of 24 patients from the UK-based study “Genetic Analysis for Personalised Medicine for Morbid Obesity” (PMMO; https://www.clinicaltrials.gov/study/NCT01365416), which recruited individuals seeking NHS treatment for severe obesity. These 24 patients underwent various types of MBS, with the majority undergoing RYGB (11 patients) and SG (9 patients). Blood samples and clinical data including weight among others were collected at up to seven time points (**Figure 1A**), during a pre-surgery assessment, during the surgery itself, and at follow-up visits ranging from 2 days to 2-3 years after surgery. Before surgery, 15 patients were diagnosed with T2D, 4 of whom were insulin dependent. Among patients with T2D, we defined subgroups based on the T2D remission status 1 year after surgery: 7 went into remission, while 8 did not, including the 4 insulin-dependent patients, giving a remission rate of 47%. The baseline characteristics of the study participants are shown in **Table 1**. The summarized anthropometric, clinical and biochemical parameters of patients from different subgroups at different time points are shown in **Table S1**.

**Figure 1.**
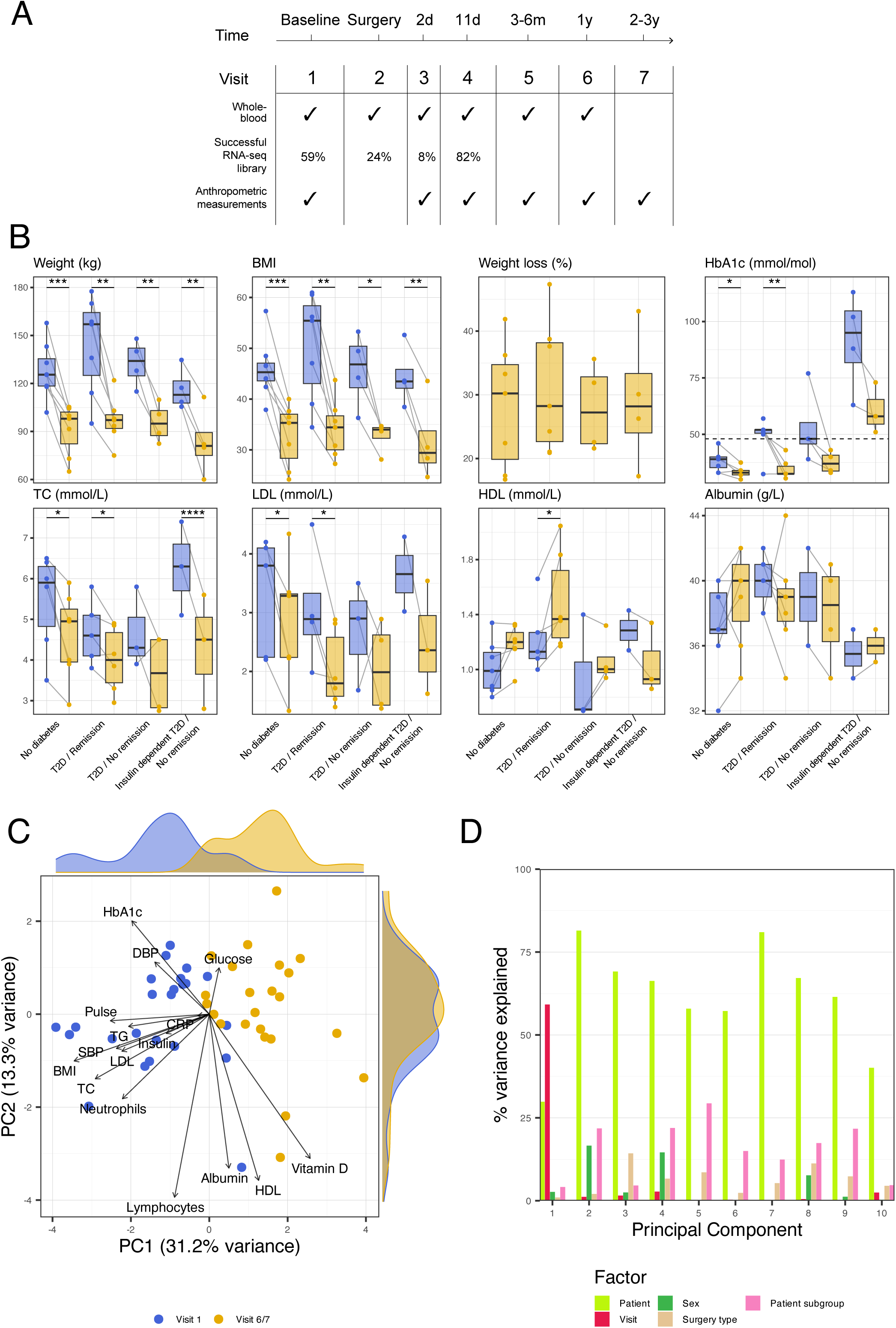
(A) Overview of the samples collection plan (d: days, m: months, y: years). The date of pre-surgery assessment visit relative to the surgery was variable across individuals (see Methods). The second row shows the percentage of patients from which RNA-seq libraries were successfully generated from collected whole blood samples, out of the 51 participants initially included. (B) Boxplots illustrating the distribution of anthropometric and biochemical measurements across patient subgroups and visits (visits 1 and averaged visits 6 and 7). Significance levels are annotated as follows: * for p ≤ 0.05; ** for p ≤ 0.01; *** for p ≤ 0.001; **** for p ≤ 0.0001. (C) Projection of the participants based on their principal component analysis (PCA) scores on principal components (PCs) 1 and 2. The visit time is indicated with colour. The margin density plots represent the distribution of participants along PCs. Contributions of variables to PCs are indicated with the orientation of arrows. (D) Fraction of variance from each of the 10 first PCs of the PCA explained by different patient metadata variables.

**Table 1.** Baseline characteristics of study participants at pre-surgery assessment visit. Parameters are summarized with mean and standard deviation across participants. The “% NA” column indicates the proportion of missing values for a given parameter across participants.

|  | Mean | SD | Counts | % NA |
| --- | --- | --- | --- | --- |
| Sex |  |  | 14 F; 10 M | 0 |
| Ethnicity |  |  | 3 Asian; 3 Black; 13 White; 5 other | 0 |
| Surgery type |  |  | 11 RYGB; 9 SG; 4 other | 0 |
| Age (years) | 46.5 | 13.3 |  | 0 |
| Weight (kg) | 132.7 | 22.3 |  | 0 |
| Height (m) | 1.7 | 0.1 |  | 0 |
| BMI (kg/m^2) | 47.2 | 7.5 |  | 0 |
| Systolic BP (mm Hg) | 141.1 | 17.2 |  | 50 |
| Diastolic BP (mm Hg) | 82.5 | 11.8 |  | 50 |
| Pulse (bpm) | 79.4 | 18.3 |  | 29.2 |
| Glucose (mmol/L) | 5.7 | 1.4 |  | 50 |
| Insulin (mIU/L) | 46.5 | 45.8 |  | 75 |
| HbA1c (mmol/mol) | 53.5 | 21.8 |  | 4.2 |
| Total Cholesterol (mmol/L) | 5.2 | 1.1 |  | 29.2 |
| LDL (mmol/L) | 3.2 | 0.9 |  | 41.7 |
| HDL (mmol/L) | 1.1 | 0.3 |  | 29.2 |
| Triglycerides (mmol/L) | 2.7 | 3.4 |  | 29.2 |
| Albumin (g/L) | 38.6 | 3 |  | 16.7 |
| Lymphocytes (10^9/L) | 2.4 | 0.9 |  | 16.7 |
| Neutrophils (10^9/L) | 6 | 1.5 |  | 20.8 |
| Vitamin D (µg/L) | 39.9 | 21.8 |  | 12.5 |

At baseline, the levels of many clinical measurements (e.g., glucose, TC, LDL levels) were elevated compared to reference ranges [22], but for most there was no significant difference across subgroups, most likely due to small sample sizes and some missing values (**Figure 1B**). Only HbA1c displayed marked differences: levels were significantly higher in insulin-dependent T2D patients than in other subgroups (p=3.6×10^-6^ compared to non-diabetic patients, p=8.6×10^-5^ compared to patients with T2D remission, and p=1×10^-3^ compared to patients without T2D remission), whereas they were much better controlled in T2D patients on oral antidiabetic drugs (not significantly different from the non-diabetic patients). The advanced DiaRem score, calculated at baseline for patients with T2D (a high score indicates a lower likelihood of T2D remission after surgery) was, as expected, significantly higher in the insulin-dependent T2D subgroup than in the other subgroups (p=1.9×10^-5^; **Figure S1**).

Next, we focused on the surgery outcomes and performed paired comparisons of phenotypic parameters between visit 1 (baseline) and visits 6 and 7 averaged (1-3 years after surgery; **Figure 1B, S1**). As expected, we found large improvements in weight and BMI in all subgroups, with percentage weight loss ranging from 16.7% to 47.4%. Overall, most blood and cardiovascular parameters were improved, reflecting the beneficial effect of MBS although not all changes were statistically significant. Noticeably, HbA1c levels decreased below the accepted remission threshold in all T2D patients on oral antidiabetic drugs, but not in insulin-dependent T2D patients. Vitamin D levels increased after surgery in all subgroups, likely due to prescribed supplementation.

A principal component analysis (PCA) was performed on 16 variables to reduce them in an unbiased way to a few uncorrelated dimensions (principal components or PCs) that capture the variability of the dataset (**Figure 1C, 1D**). This confirmed that the differences before and after surgery explained most of the variance in anthropometric and clinical variables. Most of the remaining variance was explained by patient-specific effects (**Figure 1D**). The patient subgroups defined above were not clearly separated on the first PCs, nor were other patient subgroups, e.g., defined by sex or type of surgery.

In summary, the phenotypic characterisation of our cohort confirmed the drastic effects of MBS in the months following surgery, and did not reveal any unexpected result that would undermine confidence in the transcriptome analysis described below.

### Early changes in blood transcriptome following MBS are widespread

To characterize changes in gene expression occurring shortly after MBS, RNA-seq libraries were prepared from whole blood samples collected at visit 1 (baseline) and visit 4 (median of 11 days after surgery; **Figure 1A**). A PCA using the normalized gene expression values revealed that most of the variation in the dataset was associated with patient-specific effects, but the pre- and post-surgery samples were noticeably separated on PC1 (**Figure 2A, 2B**).

**Figure 2.**
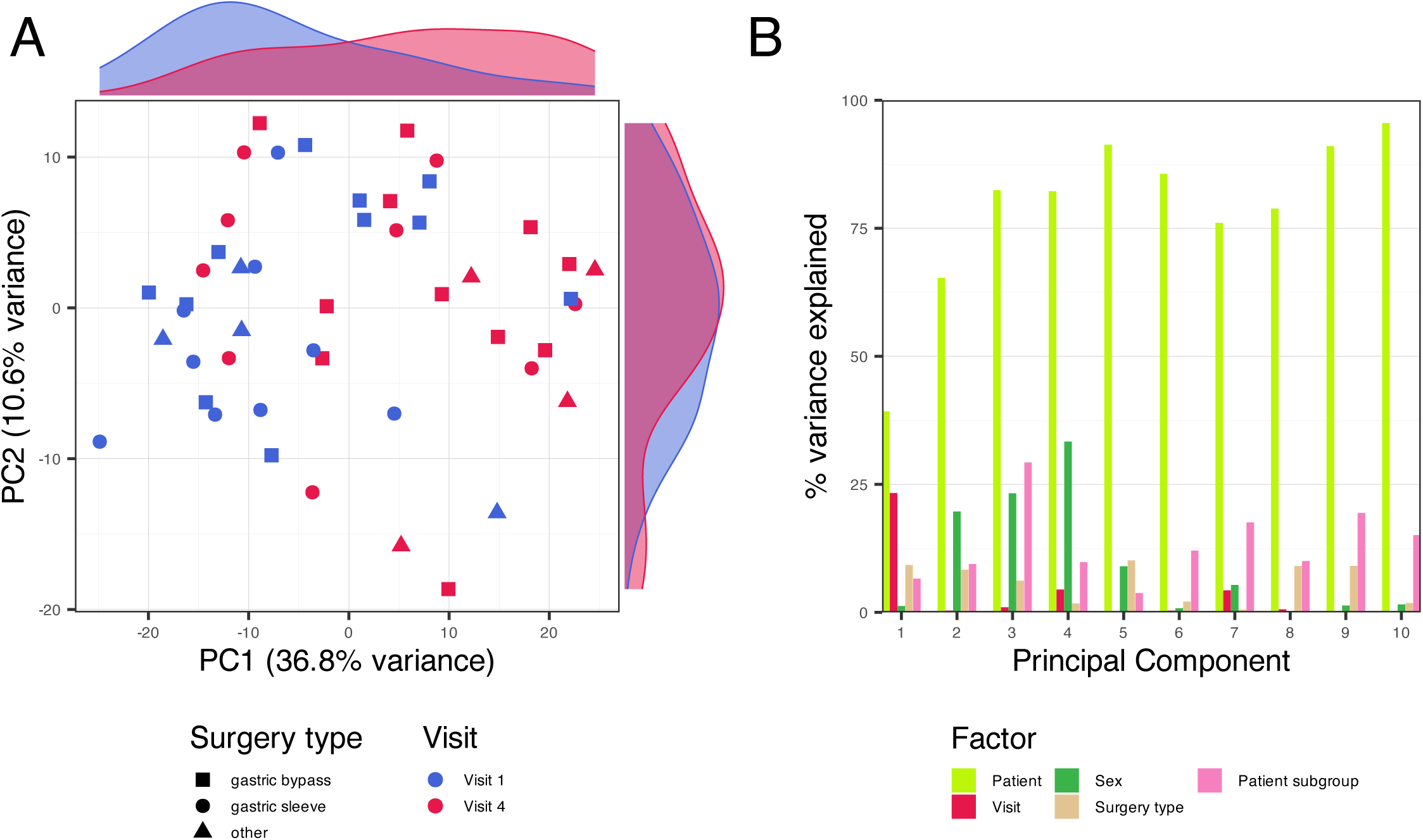
(A) Projection of the 48 RNA-seq samples based on their PCA scores on PCs 1 and 2. Legend similar to Figure 1C. (B) Fraction of variance from each of the 10 first PCs of the PCA explained by different patient metadata variables.

We performed a differential expression analysis to discover genes regulated after surgery, taking advantage of our fully-paired design to account for patient-specific effects [23]. At a false discovery rate (FDR) cutoff of 5%, 1,347 genes were differentially expressed (coloured points in **Figure 3A; Table S2**). There were twice as many down-regulated genes (884) as up-regulated genes (463). Despite the large number of differentially expressed genes, the overall fold-changes were modest (**Figure 3A, S2)**.

**Figure 3.**
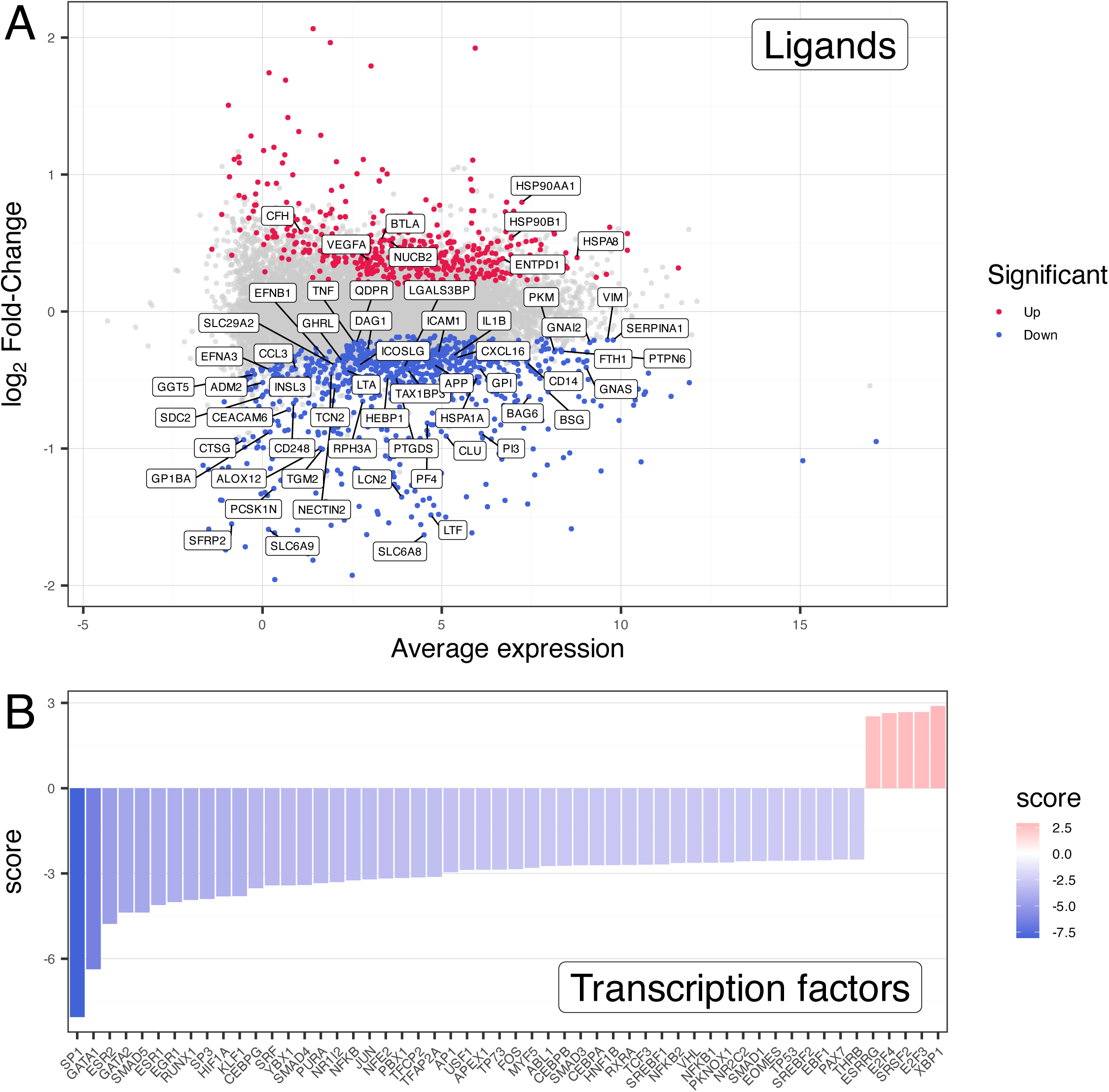
(A) MA-plot representing the log2 fold-change and average expression of genes tested for differential expression in the pre- vs. post-surgery comparison. Genes coloured in red were significantly up-regulated after surgery at an FDR of 5%, and genes coloured in blue were significantly down-regulated. Genes labelled with their symbol are coding for annotated ligands. (B) Scores of biological activity for transcription factors, inferred with the decoupleR method. A positive score reflects a higher activity after MBS.

### Expression levels of multiple cytokines, hormones and receptors are altered after MBS

To explore the differential expression results, we first focused on genes involved in signaling, in particular genes coding for ligands such as cytokines and hormones (**Figure 3A**). *TNF, LTA* and *IL1B*, coding for the key inflammatory cytokines TNFα, LTA and IL1β were down-regulated, along with several chemokine genes such as *CCL3*, *CXCL16* and *PF4*, genes involved in lymphocyte co-stimulation, such as *ICOSLG* and *NECTIN2*, and several other genes involved in immune regulation (e.g., *CEACAM6*, *LCN2*, *LTF* and *CTSG*, *CD14, GP1BA, PTPN6*). Some molecular chaperones known to be involved in the regulation of the inflammatory response, were differentially expressed, including *HSPA1A* (down-regulated), *HSP90B1*, *HSPA8* and *HSP90AA1* (up-regulated). Focusing on genes coding for hormone and cytokine receptors (**Figure S2A**), the profound rewiring of the innate and adaptive immune system was confirmed by the up-regulation of key regulators of the immune response, such as *CCR2, CCR9, CD28, CSF3R, ICOS, IL1RL1, IL7R, JAML, TLR6, TNFRSF10B* and *TNFRSF17*, and down-regulation of *ACKR1, CD40, CXCR5, FCER2, GPR84, KIR2DL3, KIR3DL1, KIR3DL2, LAMP1* and *SIGLEC10*.

Several ligands and receptors related to obesity and diabetes were down-regulated (**Figure 3A, S2A**): These included *GHRL*, coding for the appetite-regulating hormone ghrelin; *PCSK1N* encoding the protein proSAAS, a precursor of the neuropeptides PEN and bigLEN involved in appetite regulation [24]; *ADM2*, encoding the hormone adrenomedullin-2/intermedin, which affects feeding behaviour in mice [25]; *SLC6A8* encoding the sodium- and chloride-dependent creatine transporter 1, whose expression level in adipose tissue positively correlates with insulin sensitivity [26, 27]; *SLC6A9* encoding the glycine transporter 1, whose inhibition improves metabolic homeostasis in diabetes and obesity [28]; *CD248* encoding the protein endosialin, which promotes insulin resistance [29]; *SERPINA1* encoding the protein α-1-antitrypsin, which enhances insulin secretion [30]; *PKM* involved in glycolysis; *PTGDS* involved in the insulin-stimulated translocation of the glucose transporter GLUT4 [31, 32]; *CLU* encoding the protein clusterin, which is associated with adipose tissue insulin resistance [33]; *TGM2,* usually overexpressed in diabetes [34]; *ADIPOR1*, encoding the receptor for the ADIPOQ hormone secreted by adipocytes, which regulates glucose and lipid metabolism; and *LSR* encoding the lipolysis-stimulated lipoprotein receptor (or Angulin-1), which regulates triglyceride levels in the blood [35].

Receptors specific to erythrocyte were also down-regulated, such as *EPOR*, encoding the erythropoietin receptor expressed at the surface of erythroid cells; *KEL* encoding the Kell blood group glycoprotein; and *AQP1*, a component of the ankyrin-1 complex; as well as receptors involved in platelet aggregation (*GP6, ITGA2B, TBXA2R*)

### Uncovering transcription factors behind the transcriptional response to MBS

Using a transcription factor (TF) activity inference analysis, we next aimed to identify master regulators of the gene expression changes after MBS (see Methods). The activity of 54 TFs was inferred to be significantly affected (**Figure 3B**). Key regulators of the immune and inflammatory responses, such as NF-κB and AP-1, were less active after surgery, as were other immune regulators such as the CCAAT/enhancer-binding proteins (CEBPA, CEBPB and CEBPG), EBF1, EGR1, EOMES, RUNX1, SP1, THRB, and TP53.

Perhaps related to the chronic hypoxic state associated with obesity and diabetes [36], HIF1A, a master regulator of the response to hypoxia and its interactor VHL, were less active. The hypoxic response is known to trigger the transcription of multiple target genes including erythropoietin, glucose transporters, glycolytic enzymes or vascular endothelial growth factor. Other TFs known to be involved in the regulation of genes involved in glucose and lipid metabolism, such as SREBF1, SREBF2, CEBPA were also less active. XBP1, which was more active after surgery, regulates the terminal differentiation of B cells to plasma cells [37], but is also involved in glucose homeostasis and lipid metabolism regulation [38].

TFs involved in erythrocyte development (GATA1, KLF1, NR2C2, TFCP2) and muscle differentiation and regeneration (MYF5, PBX1, SMAD4, PKNOX1) were less active.

The TFs E2F3 and E2F4, known to be involved in cell cycle regulation, were more active after surgery. The orphan nuclear oestrogen related receptor gamma (ESRRG) was more active, but the oestrogen receptors alpha and beta (ESR1 and ESR2) and other nuclear receptors (NR1I2 and RXRA) were less active after surgery.

### Functional protein networks affected by MBS

Using the StringDB database [39], we organized the differentially expressed genes into a network reflecting functional associations, mostly built from known protein-protein interactions (**Figure S3**). Some clusters of interacting proteins encoded by up-regulated genes were involved in immune regulation, in particular direct interactions between B and T lymphocytes. A large cluster of proteins encoded by down-regulated genes included the subunits of the NF-κB complex, its upstream regulators and its downstream targets, and another cluster included proteins related to neutrophil cytotoxic activity.

Other clusters of proteins encoded by down-regulated genes were related to the energy metabolism of cells, in particular glycolysis, gluconeogenesis or insulin signaling. A tight cluster was composed of proteins encoded by the mitochondrial genome (all genes from the mitochondrial genome were down-regulated in our results, and 23 were statistically significant).

Finally, several clusters of proteins encoded by down-regulated genes were related to erythrocyte differentiation and function, with a cluster notably including multiple haemoglobin subunits.

We provide a more detailed description of these functional associations between differentially expressed genes in **Supplementary Text**.

### Pathway-level expression changes after MBS point at a down-regulation of inflammatory, hormone signaling and erythrocyte-related pathways

We performed a gene set enrichment analysis to identify pathways that were enriched for genes differentially expressed after MBS. We focused on pathways from the Reactome database [40], and found that 111 pathways were significant at an FDR of 5% (**Table S3**). The up-regulated pathways were related to chaperone activation (“ATF6 (ATF6-alpha) activates chaperones” pathway) and DNA repair (“Resolution of D-Loop Structures” pathway).

Most of the significant pathways were, however, down-regulated. Multiple pathways share some of their genes, so to ease exploration of the partially redundant results, we visualized the significant pathways as a network connecting pathways sharing at least 10% of their genes (**Figure 4)**. Three large clusters of interconnected down-regulated pathways were apparent: the first one was related to immune response to infection and NF-κB pathway activation. The second was related to hormone signaling (in particular glucagon signaling) and regulation of insulin secretion, involving multiple G protein-coupled receptors (GPCRs). Finally, the third cluster was related to the cytoskeletal network, involving actin and tubulin genes, genes from the cell adhesion molecule L1 family, spectrins, and members of the ankyrin complex, the latter being involved in the stability and shape of the erythrocyte membrane [41]. Other down-regulated pathways were related to other aspects of erythrocyte function, such as “Heme biosynthesis” or “Erythrocytes take up carbon dioxide and release oxygen”.

**Figure 4.**
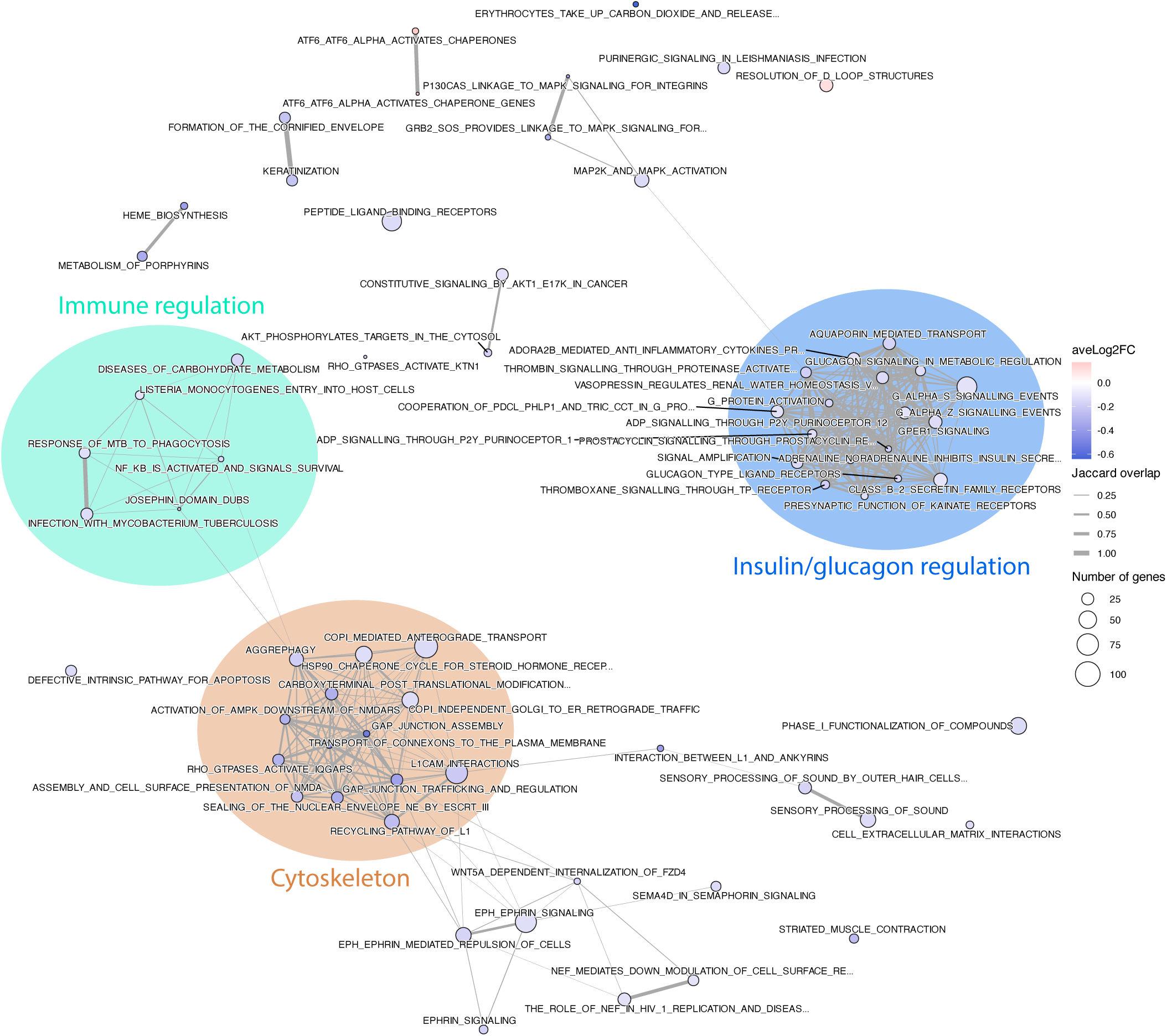
Enrichment map build from gene set enrichment analysis results using the Reactome subset of canonical pathways from the MSigDB curated gene sets (C2) collection. Only pathways significant at an FDR of 5%, including less than 100 genes and showing an absolute average log2 fold-change above 0.1 were included in the map (the full results list is shown in Table S3). The overlap of genes across pathways was calculated using the Jaccard coefficient, and an edge was drawn between pathways for a minimum coefficient of 0.1, and increasing edge width is used for higher coefficient. The number of genes mapped to each pathway is shown with the circle area, while the average log2 fold-change is shown with the circle colour.

The significant pathways obtained when using the Wikipathways [42] or Gene Ontology [43] databases were largely consistent with the results from the Reactome database (**Figure S4; Tables S4, S5; Supplementary Text**).

### MBS induces changes in abundance of erythrocytes and immune subsets

Since no quantification of cells was performed on the blood samples collected at visit 4, we inferred changes in abundances indirectly from the transcriptome profiles. We used a cell type enrichment analysis approach [44] that uses reference transcriptome signatures of pure blood cell types, and tested for differences pre- and post-surgery (**Figure 5; Table S6**). At an FDR cutoff of 5%, CD8+ Tcm and Th2 cells were inferred to be more abundant after surgery, and at a slightly higher false discovery rates, there was evidence for increased abundance of CD8+ T cells (FDR=7.5%), plasma cells (FDR=13%) and CD4+ memory T cells (FDR=19%).

**Figure 5.**
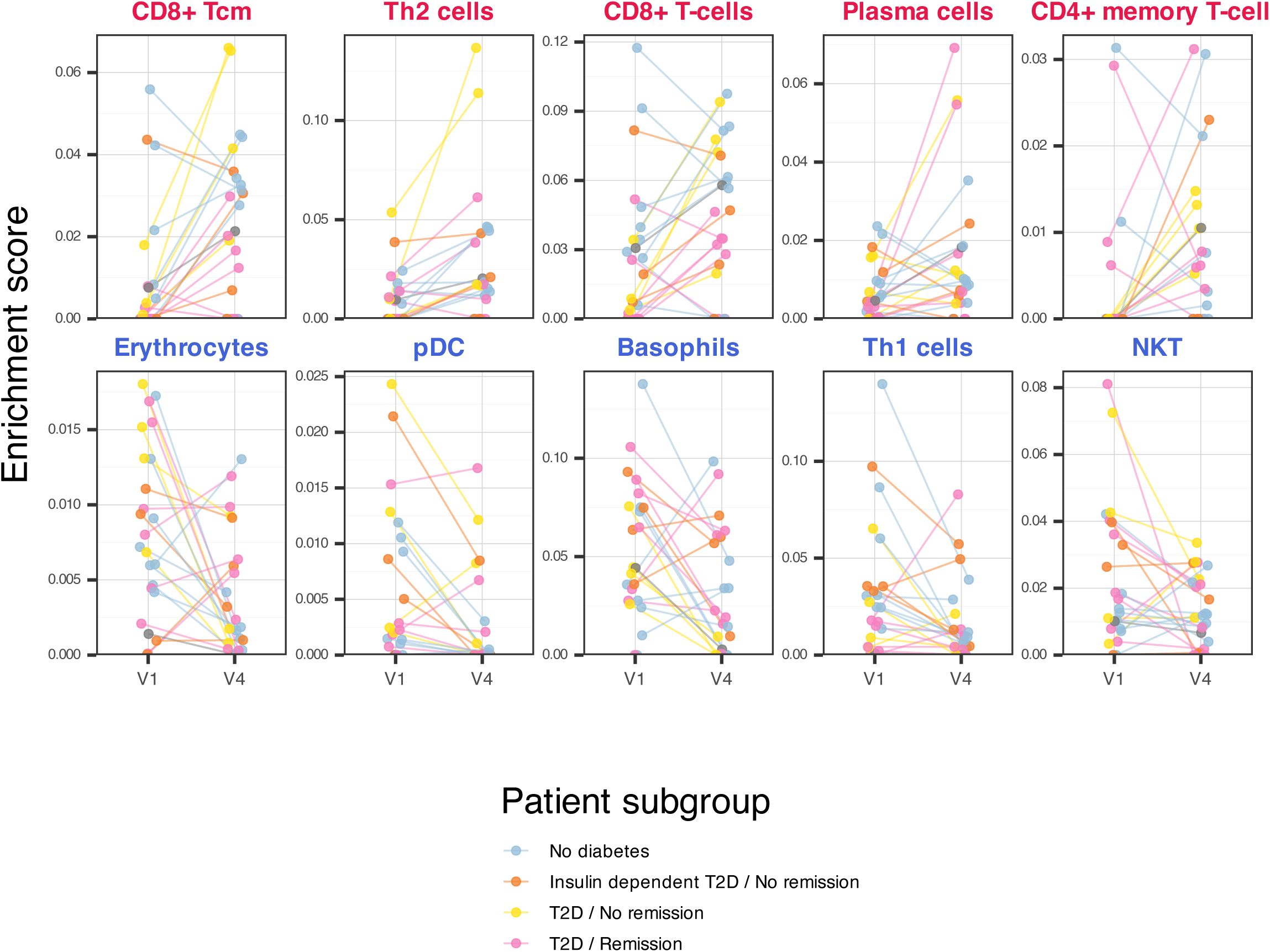
Cell-type enrichment scores for the different cell types across samples, stratified by visit. Significant cell types are shown (at an FDR=20% cutoff). Cell types are ordered by significance, with cell types inferred to be more abundant after surgery in top row and less abundant in bottom row. Lines connect matched samples from each patient. Dots and lines colour represent the patient subgroup based on diabetes status before surgery and remission status 1 year after surgery.

Conversely, erythrocytes were inferred to be less abundant after surgery (FDR=4.8%), along with pDCs (FDR=5.5%), basophils (FDR=5.5%), Th1 cells (FDR=11%) and NKT cells (FDR=19%).

The decreased abundance of Th1 helper cells and the increased abundance of Th2 helper cells, together with the increased abundance of plasma cells, prompted us to examine changes in the isotypes of immunoglobulins produced by B cells, a process modulated by cytokines produced by T cells. We focused on the differential expression patterns of genes encoding the constant region of immunoglobulin heavy chains, which determines the isotype. The genes *IGHA1* and *IGHA2*, encoding the IgA isotype produced by plasma cells of the gut-associated lymphoid tissue, were up-regulated, as was the gene *JCHAIN* (**Figure S2B**), which produces the protein that links two IgA antibodies in a dimeric form. The genes *IGHG1*, *IGHG2*, *IGHG4*, coding for the IgG isotype, which is increased in circulation in obesity and T2D-related inflammation [9], were down-regulated. The gene *IGHD,* coding for the IgD isotype, expressed on the surface of naive B cells as they leave the bone marrow, was also down-regulated.

### MBS reverses transcriptome changes associated with obesity, insulin resistance and T2D

To put the differential expression results in a broader context, we took advantage of published transcriptomic studies characterizing whole blood samples using microarrays or RNA-seq in the context of obesity, reduced insulin sensitivity and T2D. Due to patient privacy concerns, raw data from these studies were not available, thus we relied on the differential expression analysis results provided by the authors. One study investigated transcriptome changes associated with BMI using 988 individuals from the KORA F4 population-based cohort [45]. We observed a strong negative correlation (Pearson’s correlation coefficient R=-0.39, calculated across 11,719 genes common to both datasets) between the effect size estimates (beta coefficient) and the pre- vs. post-surgery fold-changes observed in our dataset (**Figure 6A**). Focusing on selected sets of genes among the most down-regulated after MBS (defined from the StringDB network analysis, see Methods; **Supplementary Text; Figure S3**), genes related to erythrocyte function, haemoglobin genes, as well as neutrophil activity genes were clearly up-regulated with increased BMI.

**Figure 6.**
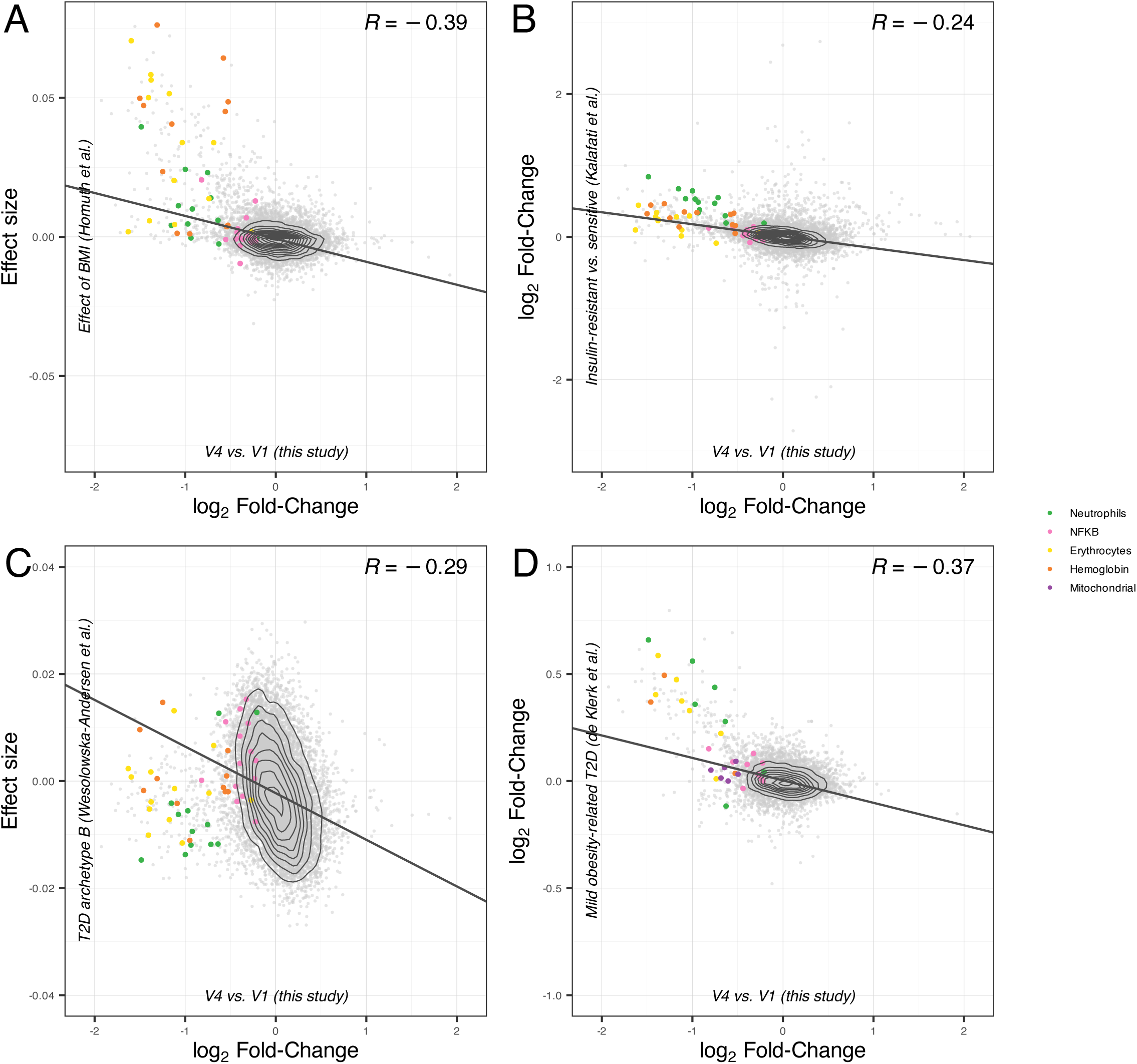
(A) Comparison of log2 fold-changes between our dataset (x-axis) and the effect size of expression changes associated with BMI in the Homuth *et al.* study (y-axis). The Pearson correlation coefficient is indicated on the top right of the plot. Coloured points represent genes belonging to gene sets of interest, defined using a StringDB interaction analysis (see Methods). (B) Similar, but the log2 fold-changes in expression between the insulin-resistant and insulin-sensitive groups in the Kalafati *et al.* study is shown on the y-axis. (C) Similar, but the effect size of expression changes associated with the scores of the T2D archetype B in the Wesolowska-Andersen *et al.* study is shown on the y-axis. (D) Similar, but the log2 fold-changes in expression between the mild obesity-related T2D subgroup and other T2D subgroups in the de Klerk *et al.* study is shown on the y-axis.

Another study compared insulin-resistant to insulin-sensitive individuals using 157 participants from the CODAM cohort, which included individuals at increased risk of T2D and cardiovascular disease [46]. Overall, gene expression changes associated with insulin-resistance were negatively correlated with those observed after MBS (R=-0.24, across 12,314 genes; **Figure 6B**), with again erythrocyte and haemoglobin genes, and neutrophil activity genes standing out in the upper-left quadrant, as up-regulated in insulin-resistant individuals.

To explore our results in the context of T2D, we turned to studies that explored the heterogeneity of this condition by stratifying patients into subgroups. A study on 726 patients with T2D from the IMI-DIRECT cohort [47] delineated four phenotypic groups (“archetypes”) using 32 clinical variables, and identified the transcriptomic signature of each phenotypic group by fitting a linear regression to the archetype scores. The transcriptome changes after MBS were most strongly correlated to the signature defining archetype B, which corresponds to patients with an obese, insulin-sensitive and relatively healthy metabolic phenotype (R=-0.29, across 12,474 genes; **Figure 6C**; correlation coefficients were R=0.034 for archetype A, R=0.18 for archetype C, and R=-0.023 for archetype D). This time, only the NF-κB activation gene set stood out with a distinctive up-regulation trend in T2D archetype B.

Another study on 400 patients with T2D from the Hoorn DCS cohort [48] defined five subgroups using the patients’ age, BMI, HbA1c, C-peptide and HDL-cholesterol levels. The “mild obesity-related diabetes” subgroup, characterized by high BMI, low age and relatively lower levels of HbA1c and HDL-cholesterol compared to other T2D subgroups, displayed the most distinct profile at the transcriptome level. We observed a strong negative correlation between transcriptome changes after MBS and those characterizing this subgroup (R=-0.37, across 7,847 genes; **Figure 6D**). All selected gene sets were overall up-regulated (as well as genes from the mitochondrial genome which were not included in the previous studies).

In summary, the transcriptome changes observed after MBS seemed overall to be opposite to those observed in obesity, insulin resistance and obesity-related T2D. Erythrocyte- and neutrophil-related genes were among the gene sets that clearly illustrated this trend.

### Comparison to long-term transcriptomic studies suggests that early transcriptome changes after MBS persist through time

Lastly, we compared our results to published transcriptomic studies that characterized the effect of MBS in the blood of patients with obesity. We found three published transcriptomic studies on whole blood or PBMCs (peripheral blood mononuclear cells), but compared to our study all had longer follow-up time points post-surgery (1 month to 1 year). For these studies, we had access to microarray/RNA-seq measurements, which allowed a complete reanalysis of the datasets using the same paired methodology used for our dataset (see Methods).

A first study sampled PBMCs from 10 patients with obesity, before and 1 month after laparoscopic sleeve gastrectomy [49]. In our reanalysis, at an FDR of 5%, 291 genes were up-regulated and 170 down-regulated 1 month after surgery. There was a positive but only weak correlation between the log-fold changes in this study and those from our study (R=0.13, across 14,114 genes; **Figure 7A**), but for the neutrophil activity and haemoglobin genes a strong down-regulation trend was commonly observed across datasets. The mitochondrial and erythrocyte-related genes were only slightly, but consistently down-regulated, whereas the NF-κB activation genes were up-regulated in this dataset.

**Figure 7.**
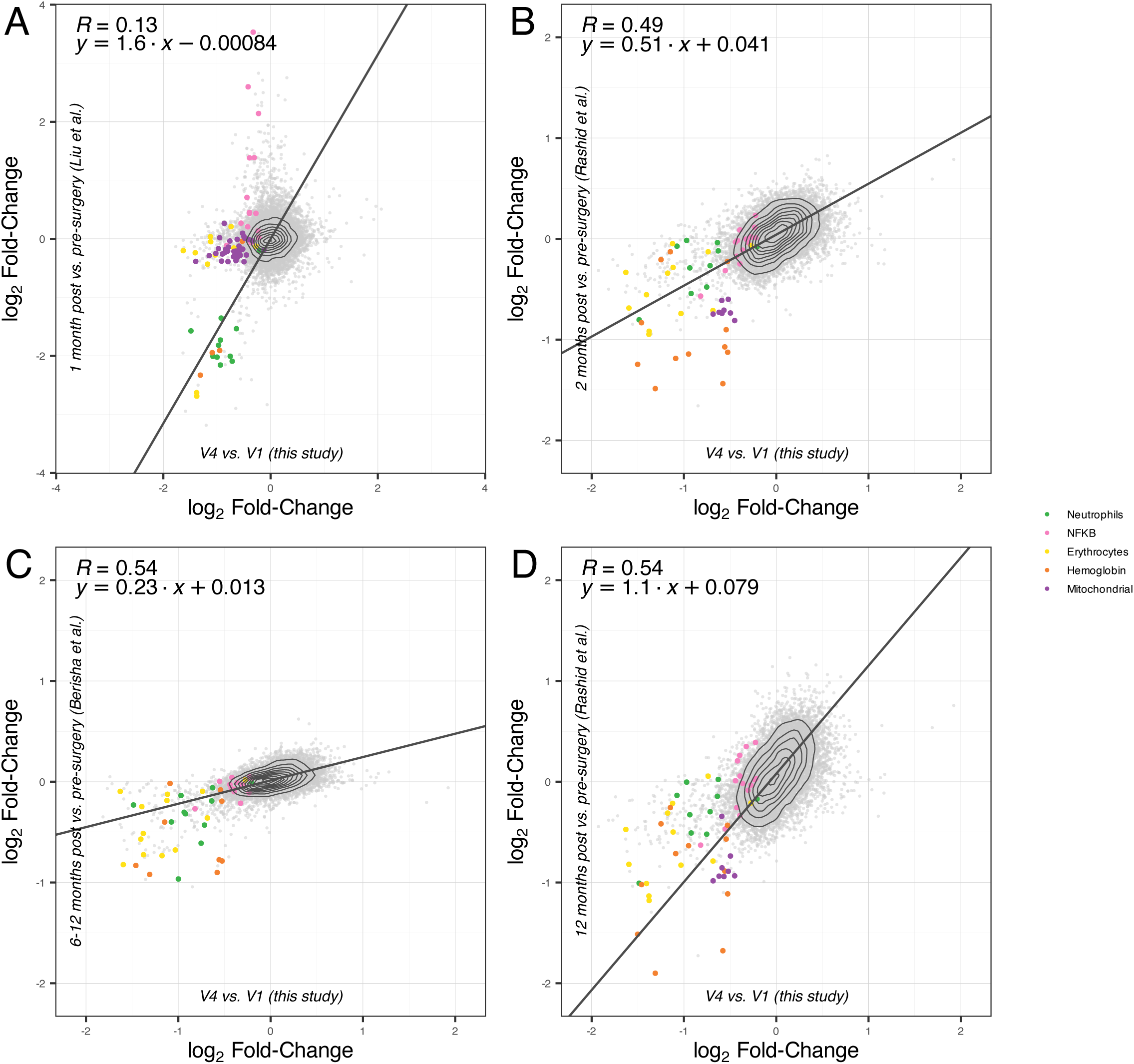
(A) Comparison of log2 fold-changes between our dataset (x-axis) and the log2 fold-changes from the reanalysis of the Liu *et al.* dataset (y-axis). X- and y-axis scales are identical. The Pearson correlation coefficient is indicated on the top left of the plot, along with the regression line slope and intercept coefficients. Coloured points represent genes belonging to gene sets of interest, defined using a StringDB interaction analysis (see Methods). (B) Similar, but the log2 fold-changes from the reanalysis of the Rashid *et al.* dataset are shown on the y-axis (pre- vs. 2 months post-MBS comparison). (C) Similar, but the log2 fold-changes from the reanalysis of the Berisha *et al.* dataset are shown on the y-axis. (D) Similar, but the log2 fold-changes from the reanalysis of the Rashid *et al.* dataset are shown on the y-axis (pre- vs. 1-year post-MBS comparison).

A second multi-centre study compared PBMC samples from 26 patients with obesity and T2D before, 2 months and 12 months after RYGB, SG, biliopancreatic diversion without duodenal switch, or laparoscopic mini gastric bypass procedures [50]. Sixteen patients (62%) were in remission 12 months after surgery. After 2 months, no gene was significantly regulated at an FDR of 5%, but we nonetheless observed a strong correlation of the log-fold changes in both datasets (R=0.49, across 11,593 genes; **Figure 7B).** The regression line slope was 0.51, which may indicate that the changes were attenuated in this dataset after 2 months compared to our dataset. All selected gene sets were down-regulated in both datasets, with the exception of NF-κB activation genes. Surprisingly after 12 months, in the same cohort, the fold changes correlated better with our dataset (R=0.54; **Figure 7D**), and the regression line slope of 1.1 indicated comparable effect sizes to our dataset. The NF-κB activation gene set was slightly up-regulated at this time point.

Last, a study compared whole blood samples from 11 patients with obesity and T2D before and 6-12 months after RYGB or SG procedures [51]. After surgery, eight patients (73%) presented normal fasting plasma glucose levels. At an FDR of 5%, only three genes were significantly differentially regulated (*LCN2, CEACAM8, CAMP,* all down-regulated). We observed again a strong correlation of the log-fold changes in both datasets (R=0.54, across 10,648 genes; **Figure 7C)**, but the regression line slope was 0.23, indicating attenuated changes in this dataset. All selected gene sets were down-regulated here, except NF-κB activation genes.

In summary, the transcriptome changes observed in published studies using longer follow-up time points after MBS (1-12 months post-surgery) were positively correlated with those observed in our study, suggesting that many changes observed days after MBS could be maintained in the long term. One notable exception to this trend was the NF-κB activation gene set, down-regulated a few days after MBS in our dataset, and up-regulated in 2 out of the 3 datasets reanalysed here.

## Discussion

The results of our cohort study provide the first report of gene expression changes in the blood of patients with obesity occurring early after MBS. To our knowledge, all previously published transcriptomic studies sampled operated patients at a later time point, typically 6 to 12 months post-surgery, providing results that are confounded by the effect of the major weight loss typically following MBS [52]. Insight into the early effects of MBS itself is of particular interest, as this surgery is associated with health benefits that are already apparent by the time of the patients’ hospital discharge, such as improved glycaemic control, and even remission in some patients with T2D. We used a rigorous analytical approach taking advantage of matched pre- and post-surgery samples to control for patient-specific effects. This allowed us to achieve good statistical power with a sample size of only 24 patients, while maintaining strict control of false discovery rate.

### Towards a resolution of the obesity-related inflammation

We found that the majority of early transcriptome changes induced by MBS were immune-related. With regard to the adaptive immune system, key genes involved in B and T cell responses, and in the crosstalk between these two cell types were affected. The ratio of Th1 to Th2 helper cells was decreased, reflecting a shift from a pro-inflammatory to a more anti-inflammatory response. This shift is associated with decreased activity of the NF-κB pathway, changes in the profile of cytokines in the bloodstream. In particular, we found that the genes coding for the key pro-inflammatory cytokines TNFα, LTA and IL1β were significantly down-regulated after MBS. These results are consistent with the literature, where several studies have reported a systemic chronic inflammatory phenotype in obesity and in obesity-related insulin resistance and T2D [4, 8, 9, 53–55], which is durably reversed by MBS [12, 19, 56, 57].

MBS also affected the innate immune system: In particular, we observed differential expression of genes related to neutrophil cytotoxic activity. Neutrophil numbers are known to be elevated in obesity and obesity-related metabolic dysregulation [58, 59], and their transcriptome profile was shown to be altered in patients with T2D [60, 61]. A reduction in the number of circulating neutrophils was previously reported in the months following MBS [51, 62], but we did not detect significant changes in neutrophil abundance in our deconvolution analysis. Our results are more consistent with a change in the activation state of neutrophils, similar to a previous report of decreased expression of neutrophil inflammatory genes in adipose tissue after MBS [63]. However, it is not possible to infer from our data whether this change in neutrophil activity is directly caused by MBS or a downstream consequence of the changes affecting the adaptive immune response discussed above.

### Genes involved in metabolic activity are regulated

Differential expression of genes involved in metabolic pathways, in particular related to glucagon and insulin signaling, highlights the rapid effects of MBS on glycolysis and gluconeogenesis. Metabolic pathways are known to interfere with inflammatory pathways in the context of obesity; in particular, the NF-κB pathway [2], which is responsible for low-grade inflammation in metabolic tissues, is thought to be critical in the development of insulin resistance. The changes in metabolic activity genes could be caused by the resolution of the inflammation, but it is unclear whether such causal changes are likely to be observed only a few days after surgery.

MBS also has direct effects on hormone secretion, in particular gut hormones involved in the regulation of digestion, glucose control and appetite [20]. In our data, we observed down-regulation of key hormones, hormone precursors and hormone receptors that regulate feeding and glucose or lipid metabolism. In particular, the down-regulation of the *GHRL* gene coding for the appetite-regulating hormone ghrelin is consistent with previous studies reporting a decrease in fasting ghrelin levels after MBS [62, 64, 65], and can mechanistically be explained by the fact that the RYGB and SG procedures resect the gastric fundus where ghrelin is produced. We hypothesize that such hormone rewiring is likely behind the rapid changes in expression of metabolic pathways observed in our dataset. However, more data is needed to assess the role that these changes have in the improvement of glycaemic control. Of note there was no detectable expression in our blood dataset of the pro-glucagon gene (GCG), precursor to the GLP-1 peptide, a key hormone to the glycaemic control changes and whose analogues are an effective treatment for both obesity and T2D.

### The temporal scale of the effects of MBS on the blood transcriptome

A new and remarkable aspect of our results is that widespread transcriptome changes related to inflammation and metabolic improvement are observed only days after MBS, despite the inflammation caused by the surgery itself. Few studies have examined such early time points, and the evidence for the timing of the decrease of the inflammation has been conflicting. A meta-analysis confirmed a significant reduction of CRP levels at follow-up of less than 3 months, but there was no significant reduction of IL-6 (only significant at 6 months and longer), and TNFα (only significant at 12 months and longer)[18]. A study focusing on lymphocytes concluded that, although the cell composition returned to that of lean controls 3 months after MBS, T and B cell functionality was not fully restored [66]. Measurement of metabolic and inflammatory markers 3 and 6 months after MBS revealed that serum concentrations of the pro-inflammatory cytokines TNF-α, IL-6, IL-8 and IL-1β were decreased significantly only at the 6-month time point [19]. The study with the earliest identified follow-up time point analysed the immediate post-operative serum cytokine profile at the time of patient discharge from hospital. Some pro-inflammatory markers (TNFα and IL-8) were reduced immediately after surgery, but some (CRP and IL-6) were increased [67].

One possible explanation for the inconsistent results between studies is that the inflammation may not be completely resolved for several weeks or months. Another possibility is that there are cohort-specific effects, for example due to differences in patients’ eligibility criteria, post-operative care practices, or biological and technical differences in the sampling strategies. Last, there is the possibility of a “rebound” of the inflammation. For example, the above-cited study of the post-operative serum cytokine profile revealed that levels of TNFα, reduced immediately after surgery, then increased above baseline at a 6 months follow-up [67]. MBS mechanically restricts food intake, but also bypasses parts of the digestive system, leading to malabsorption of iron or vitamins. Such deficiencies could cause an increase in inflammation several months after surgery [68].

### Comparison to previously published long-term studies of the effects of MBS on the blood or PBMC transcriptome

Compared to our dataset, all identified studies for which data was available had later post-surgery sampling time points, ranging from one month to one year. Overall, there was a remarkably good correlation between the expression changes observed in our study and those observed in these studies, despite the differences in sample preparation, transcriptomic technologies used, and specificities of the cohorts studied.

This suggests that many of the changes observed a few days after surgery are robust and persist over a longer period of time. Importantly this also implies that many changes occurring after MBS are initiated before any major weight loss could occur.

We observed that specific gene sets behaved differently across datasets, in particular genes from the NF-κB pathway. These were clearly up-regulated after MBS in one dataset (1 month after MBS)[49], and slightly up-regulated in another (1 year after MBS)[50]. This trend suggests complex temporal patterns in the resolution of the inflammation, which should be addressed in future work sampling the same patients at multiple follow-up time points.

### Down-regulation of mitochondrial genes after MBS

Among other notable trends in our results, genes from the mitochondrial genome were down-regulated after MBS (23 being so with an FDR below 5%). Some genes from the nuclear genome with mitochondria function were also down-regulated, but for these the overall trend was not clear (e.g., there was no significant trend for the 1,422 genes annotated to the Gene Ontology term “mitochondrion”; FDR=63%, **Table S5**). Although the expression dynamics between mitochondrial and nuclear gene can differ greatly [69], this result could indicate a decrease in mitochondrial copy number after MBS.

Genes with mitochondrial function were previously reported to be up-regulated in obesity [45, 70] and in obesity-related T2D (**Figure 6**), presumably related to an increased oxidative stress [12]; and down-regulated after MBS (**Figure 7**)[71], a result that may not be surprising, since inflammation and oxidative stress are tightly connected processes. However, the expression of genes encoded by the mitochondrial genome itself were not always studied; probes targeting these genes are often missing from microarray platforms.

### Decreased erythropoiesis after MBS

In our data, we observed a strong effect of MBS surgery on genes involved in erythrocyte differentiation and function, but also on structural components of the erythrocyte cells, such as members of the ankyrin complex on the erythrocyte membrane [41]. A deconvolution analysis suggested that this pattern was caused by a decreased abundance of erythrocytes after MBS.

This could potentially be explained by blood loss during surgery, but all patients underwent laparoscopic procedures, which minimise blood loss. Iron deficiency is also commonly observed after MBS, potentially leading to anaemia [68], but we would not expect to observe this effect only days after MBS. Another explanation could be the known association of obesity with increased erythropoiesis [45, 72], possibly as a consequence of oxidative stress impacting erythrocyte survival (**Figure 6)**. The decreased abundance of erythrocytes after MBS could therefore indicate a return towards normal state. Of note, this trend is also observed in the reanalysed MBS transcriptomic studies (**Figure 7**)[49–51, 62].

### Limitations of our study

Among the limitations of our study, the pre- vs. post-surgery comparison does not only capture the effects of MBS, but also the effects of the wound healing, the intake of anaesthetics and analgesics, the patients’ forced physical inactivity and the change in diet (patients switch to a liquid food diet and reduce their overall food intake). Some patients experience dehydration due to difficulties in water intake.

The time between pre-surgery assessment (visit 1) and MBS was also variable across patients, and it is possible that the patients changed their habits in this period, contributing to the changes observed in the differential expression analysis comparing visit 1 to visit 4. To investigate this possibility, we ran a differential expression analysis incorporating the time between pre-surgery assessment and surgery as a covariate, but no significant gene was detected (not shown).

Finally, our sample size of 24 patients, although large enough to confidently identify gene expression changes induced by MBS [23], was too small to investigate with reasonable statistical power trends specific to subgroups of patients, e.g., differences between patients with and without improvement in long-term T2D status. A rigorous test for such changes requires testing for an interaction between the pre- vs. post-surgery main effect and a subgrouping variable, which has little statistical power to detect sizeable effects [73].

## Conclusion

Our transcriptome-wide analysis provides novel insights into the early molecular events occurring after MBS, a surgery typically associated with rapid improvements in glycaemic parameters in patients with obesity and T2D. We show that only days after surgery, molecular changes are widespread and point to both a reduction in chronic inflammation and to early metabolic rewiring in operated patients.

## Material and Methods

### Study design and ethical permissions

Participants were recruited as part of the “Personalised Medicine for Morbid Obesity” cohort, an on-going multi-centre observational prospective study which has recruited over 2,700 individuals since its establishment in 2011 (https://classic.clinicaltrials.gov/ct2/show/NCT01365416). The study was conducted in accordance with the ethical guidelines of the Declaration of Helsinki II, regional ethical permission for this study was provided by NRES committee London riverside (REC: 11\LO\O9356) and Brunel University London ethics committee (BREO: 26158), and written informed consent was obtained from all participants.

### Analysis of anthropometric and biochemical measurements

Individuals (n=51) included in the current study were recruited and underwent data collection between 2011 and 2017 at St. Mary’s Hospital, London, United Kingdom. Individuals were included if they were between 18 and 65 years old, had a BMI ≥40, or ≥35 kg/m^2^ with comorbidities at the time of recruitment and were pursuing bariatric surgery. Individuals were excluded if they had donated blood in the past 3 months or received or intended to receive treatment with a drug which had not been approved by the European Medicines agency at the time of recruitment. Among the subset of 24 patients retained in this study because they had paired RNA-seq libraries prepared before and after surgery, 14 were female and 10 were male, and different types of surgeries were performed: RYGB (11 patients), SG (9 patients), gastric banding (1 patient) and revisions towards more restrictive surgeries (3 patients).

Clinical data and blood samples were collected at specified time points, while most patients were in fasting conditions: During a pre-surgery assessment (visit 1; 11 to 592 days before surgery, median of 146 days across all patients), during the surgery itself (visit 2), 2 days after surgery if the patient was still at the hospital (visit 3), and during 4 follow-up visits: 8 to 22 days (median of 11 days across all patients – visit 4), 3 to 6 months (visit 5), 1 year (visit 6) and 2 to 3 years after surgery (visit 7).

Blood samples were used for the measurement of glycated haemoglobin levels (HbA1c), albumin, vitamin D, insulin and C-reactive protein (CRP) and routine laboratory analyses (low-density lipoprotein (LDL), high-density lipoprotein (HDL), total cholesterol (TC), triglycerides (TG)). Anthropometric traits (weight, height (from which was derived body mass index (BMI), sex, age, pulse, systolic and diastolic blood pressure (SBP and DBP respectively) and drug therapy (antihypertensive, GLP1 analogues, insulin intake, lipid lowering and oral antidiabetic drugs) were also reported. Weight loss was computed across the different visits compared to pre-surgery (visit 1). The sex of the patients was confirmed using the RNA-seq data, by looking at the expression level of the XIST lncRNA gene (detected above background level in female samples).

Subgroups of patients were created, based on the diabetes status during pre-surgery assessment (visit 1), and 1 year after the surgery (visit 6). Visits 5 and 7 were also checked. The ADA-defined criteria for diabetes remission were used, namely HbA1c levels < 6.5% (= 48 mmol/mol) in absence of glucose-lowering pharmacotherapy [74]. An underestimation of patients with T2D remission could occur since the medication Metformin was not always stopped following MBS in patients with metabolic improvement.

For patients with T2D, the advanced DiaRem score was calculated according to Aron-Wisnewsky *et al*. [75]. Score ranges from 0 (higher chance of remission) to 21 (lower chance) with an accepted threshold of 10 considered for remission.

A principal component analysis was performed using the *pca()* function from pcaMethods R package (version 1.92.0) and with the following options: method=“nipals” in order to manage missing values. We considered the variables BMI, HbA1c, glucose, vitamin D, SBP, DBP, pulse, lymphocytes, neutrophils, CRP, insulin, albumin, TC, HDL, LDL and TG for samples at visit 1. The values were averaged for samples at visits 6 and 7 to decrease the amount of missing data. Percentages of variance explained by sex, visit, subgroups, type of surgery and patient were estimated using the *getExplanatoryPCs()* function from the Bioconductor package scater (version 1.28.0). Testing of differences in clinical parameters values across patient subgroups at visit 1 was done with the *glht()* function from multcomp R package (version 1.4-25), using a post-hoc Tukey test. For comparisons between visits 1 and 6-7 a linear mixed-effect model was used with patient as random effect (R package nlme, version 3.1-163)

### RNA-sequencing data collection

Blood samples were collected using PAXgene Blood RNA tubes (Qiagen), and RNA extraction was performed on samples from visit 1, visit 2, visit 3 and visit 4 (**Figure 1A**). After quality control the resulting RNA was stored at -80°C for RNA-seq library preparation. Illumina library preparation and sequencing were performed at the Oxford Genomics Laboratories. Sequencing was performed for 91 RNA-seq samples across 51 patients on an Illumina Hiseq 2000 instrument to produce paired-end 92 bp-long reads. Matched samples pre- (visit 1) and post-surgery (visit 4) were available for 24 out of the 51 patients.

### RNA-sequencing data analysis

Data analysis was performed at the Bioinformatics Core Facility, Department of Biomedicine, University of Basel. Read quality was assessed with the FastQC tool (version 0.11.8). Reads were mapped to the human genome hg38 with STAR (version 2.7.9a)[76] with default parameters, except filtering out multimapping reads with more than 10 alignment locations (outFilterMultimapNmax=10) and filtering reads without evidence in the spliced junction table (outFilterType=“BySJout”). The number of reads mapped to the genome ranged from 18M to 40M reads per sample.

The *featureCounts()* function from the Bioconductor Rsubread package (version 2.0.1)[77] was used to count the number of reads (5’ ends) overlapping with the exons of each gene (Ensembl release 102) assuming an exon union model. All subsequent analyses were performed using the R software (version 4.3.1) and Bioconductor 3.18 packages [78].

A total of 14,358 genes with CPM values above 1 in at least 12 samples (1/4 of the samples), and not assigned to the pseudogene or TEC (to be experimentally confirmed) biotypes, were retained for the differential expression analysis. To control for compositional changes across samples, a cyclic loess normalization [79] was chosen as a good compromise between the scaling factors normalization, unable to correct for an observed trend between log fold-changes and the average level of expression, and a more aggressive normalization such as quantile normalization.

The sex of the patients was verified by looking at the expression level of the *XIST* lncRNA gene (detected above background level only in female samples).

A principal component analysis was performed on the normalized logCPM values from the 500 genes with the largest inter-quartile range values. The association with patient metadata and experimental factors was tested using the function *getExplanatoryPCs()* from the Bioconductor package scater (version 1.30.0).

Differential expression analysis between visit 1 and visit 4 was performed using the *voom()* function [80] from the Bioconductor package limma (version 3.58.1), by fitting an additive model accounting for the visit and patient-specific effects (and the *robust=TRUE* option in the *eBayes* function). P-values were adjusted by controlling the false discovery rate (FDR; Benjamini-Hochberg method) and genes with an FDR lower than 5% were considered significant. For the patient subgroup analyses according to sex, surgery type, or diabetes remission status, an interaction between visit and subgroup was tested.

The large number of samples allowed us to use a self-contained testing strategy for gene set enrichment analysis, which considers each pathway independently of the others and explicitly tests whether the majority of genes within a pathway are differentially expressed. This was performed with the function *fry()* from the limma package [81]. Fry is a fast alternative to *mroast()* with the parameters *set.stat=“mean”* and a very large number of rotations. Gene sets from the MSigDB Molecular Signature Database (version 2023.2.Hs)[82] were used, with a focus on the C2 Reactome [40] and Wikipathways collections [42], as well as the Gene Ontology categories (mappings obtained from http://current.geneontology.org/ with a date stamp of 2023-07-27)[43]. Gene sets containing less than 10 genes were filtered out, and those with an FDR lower than 5% were considered significant. For visualization of the enrichment network, a Jaccard coefficient was used to quantify the gene overlap across gene sets. Gene sets with not more than 200 genes were plotted, and connected by an edge if their overlap was above 0.2.

Protein-protein interactions networks for differentially expressed genes were created with StringDB (version 12)[39]. The full STRING network was visualized, displaying all interaction sources except co-expression that had highest confidence scores (>=0.9). Only query proteins were shown, hiding disconnected nodes in the network, and without any first shell or second shell interactors. A Markov clustering (MCL) with inflation parameter set to 1.5 was used to delineate clusters of interacting proteins. Among the largest clusters, four were selected to highlight specific sets of genes in the comparisons to published datasets: Neutrophil activity genes (*AZU1, BPI, CEACAM6, CEACAM8, CES1, CTSG, DEFA1B, DEFA4, ELANE, LTF, MPO, PRTN3, SERPINA1*), erythrocyte membrane and cytoskeleton genes (*ADD1, ANK1, CA2, DMTN, EPB42, GYPB, GYPC, MPP1, RHD, SLC4A1, SPTA1, SPTB, TMOD1*), haemoglobin and erythrocyte function genes (*AHSP, CYB5R3, HBA1, HBA2, HBE1, HBM, HBQ1*, to which we added all other haemoglobin subunits), and NF-κB activation genes (*BCL3, CCL3, CD40, CXCR5, ICAM1, ICOSLG, IL1B, LTA, NFKB1, NFKB2, NFKBIB, RELA, RELB, SHARPIN, TNF*). A last gene set was made of all genes from the mitochondrial genome.

The list of annotated ligands and receptors was compiled using multiple sources. From the R package CellChat (version 2.1.2)[83], only interactions annotated as “Secreted Signaling” were considered. Interactions from the CellTalkDB database [84] were obtained from https://github.com/ZJUFanLab/CellTalkDB (file *human_lr_pair.rds* last updated on Aug 31, 2020). Last, interactions from the CellPhoneDB database [85] were obtained from https://github.com/ventolab/cellphonedb-data (Release v5, last update on Sept 11, 2023) and only interactions with directionality “Ligand-Receptor” were considered.

Transcription factors activity was inferred using the decoupleR package (version 2.9.7)[86]. As input we used the CollecTRI collection, including a curated collection of TFs and their targets compiled from multiple resources, accessed using the OmnipathR package (version 3.13.18)[87]. TF complexes were not split into subunits. The TF activity scores were obtained with the Univariate Linear Model method, using as input the *t*-values of the differential expression test between visits 1 and 4. TFs with an FDR below 10% were considered significant.

### Deconvolution analysis

Inference of abundance of cell types from blood samples based on the expression profiles measured by RNA-seq was performed using the xCell tool [44], a gene signatures-based method that can be used to infer up to 64 immune and stromal cell types. The xCell R package (version 1.1.0) was obtained from https://github.com/dviraran/xCell and the *xCellAnalysis()* function was used on the log-transformed RPKM values to perform the 3 analysis steps (rawEnrichmentAnalysis, transformScores, and spillOver). A subset of 33 cell types was considered, excluding cell types not expected to be present in blood samples, and cell types which yielded enrichment scores close to 0 in the vast majority of samples. Testing of significance abundance changes between visit 1 and visit 4 samples was done using the limma package (version 3.58.1) using the options *trend=TRUE* and *robust=TRUE* in the *eBayes* function, and accounting for patient-specific effects in the model. P-values were adjusted by controlling the false discovery rate (FDR; Benjamini-Hochberg method).

### Comparison to published datasets

Results from the Homuth *et al*. study [45], i.e., effect sizes from a linear regression testing the association of gene expression levels in the whole-blood and BMI in the KORA F4 cohort, were obtained from Table S34 of the article (“basic model”, adjusting for age, sex, technical covariates, and blood cell counts). The Bioconductor package illuminaHumanv3.db (version 1.26) was used for gene annotation of microarray probes from the Illumina HumanHT-12 v3 beadchip. Effect sizes of probes matching the same Ensembl gene were averaged.

Results from the Kalafati *et al.* study [46], i.e., log fold-changes from the differential expression analysis comparing insulin-resistant to insulin-sensitive individuals from the CODAM cohort, were obtained by email request to the authors (“model 1”, adjusting for sex, BMI and age).

Results from the Wesolowska-Andersen *et al*. study [47], i.e., effect sizes from a linear regression testing the association of gene expression levels in the whole blood to the patients’ scores on each archetype, were obtained from Table S5F of the article (results adjusted for age, sex and recruitment centre).

Results from the de Klerk *et al.* study [48], i.e., log fold-changes from the differential expression analysis comparing the MOD T2D subgroup to other subgroups, were obtained from ESM Table 5 of the article (results adjusted for the first three PCs of the blood cell fractions and technical covariates).

Raw data (.fastq) files from the Liu *et al.* study [49] were obtained from the ENA database (BioProject ID PRJNA861382). Read mapping and gene counting was performed as above-described for our dataset using STAR and featureCounts. The differential expression analysis to compare samples pre- to post-surgery was performed similarly to our dataset, using the limma-voom method. For the fitting of a regression line in the comparison of log-fold changes across MBS datasets, a total least squares (orthogonal) regression was performed using the *prcomp()* function.

Raw data files (.CEL files) from the Rashid *et al.* study [50] were obtained by email request to the authors and are now available on GEO (accession GSE271700). The Bioconductor packages affy (version 1.80) and hgu133plus2.db (version 3.13) were used to load the raw data and for gene annotation of microarray probes from the Affymetrix Gene Chip Human Genome U133 Plus 2.0 array. Probes matching the same Ensembl gene were averaged. Gene expression levels were normalized using a cyclic loess normalization [79]. The differential expression analysis to compare samples before and after surgery was performed similarly to our RNA-seq dataset, using the limma package by fitting an additive model accounting for the visit and patient-specific effects (using the *trend=TRUE* and *robust=TRUE* options in the *eBayes()* function). P-values were adjusted by controlling the false discovery rate (FDR; Benjamini-Hochberg method).

Microarray data from the Berisha *et al.* study [51] were obtained from GEO using the GEOquery package (version 2.70; accession GSE19790). The Bioconductor package illuminaHumanv2.db (version 1.26) was used for gene annotation of microarray probes from the Illumina human-6 v2.0 expression beadchip. Probes matching the same Ensembl gene were averaged. The differential expression analysis was performed similarly to the Rashid *et al.* dataset.

---

Table S1 Characteristics of study participants grouped by T2D status subgroup, across visits. Parameters are summarized with mean and standard deviation across participants in a subgroup. “NA” values indicate missing values for a parameter across participants in a subgroup. “(+/-NA)” indicates that the standard deviation could not be calculated, i.e., that only one participant had non-missing value.

Table S2 Results from the differential expression analysis comparing visit 1 and visit 4, and accounting for patient-specific effects. The “logFC” column displays the log2-transformed fold change estimate (positive for genes up-regulated at visit 4 compared to visit 1). The “AveExpr” column displays the average normalized expression level calculated across all samples. The “adj.P.Val” displays the p-value adjusted for multiple testing (FDR).

Table S3 Results from the enrichment analysis using pathways from the Reactome database. The “NGenes” column indicates the number of genes in each pathway that were included in our differential expression analysis. The “aveLog2FC” column indicates the average log2 fold-change across genes in each pathway.

Table S4 Similar to Table S3 for the enrichment analysis using pathways from the Wikipathways database.

Table S5 Similar to Table S3 for the enrichment analysis using gene sets from the Gene Ontology database. Molecular Function MF, Cellular Component CC, and Biological Process BP ontologies were used.

Table S6 Results from the cell type differential abundance analysis comparing visit 1 and visit 4, and accounting for patient-specific effects, based on the xCell enrichment scores in each sample.

**Figure S1.**
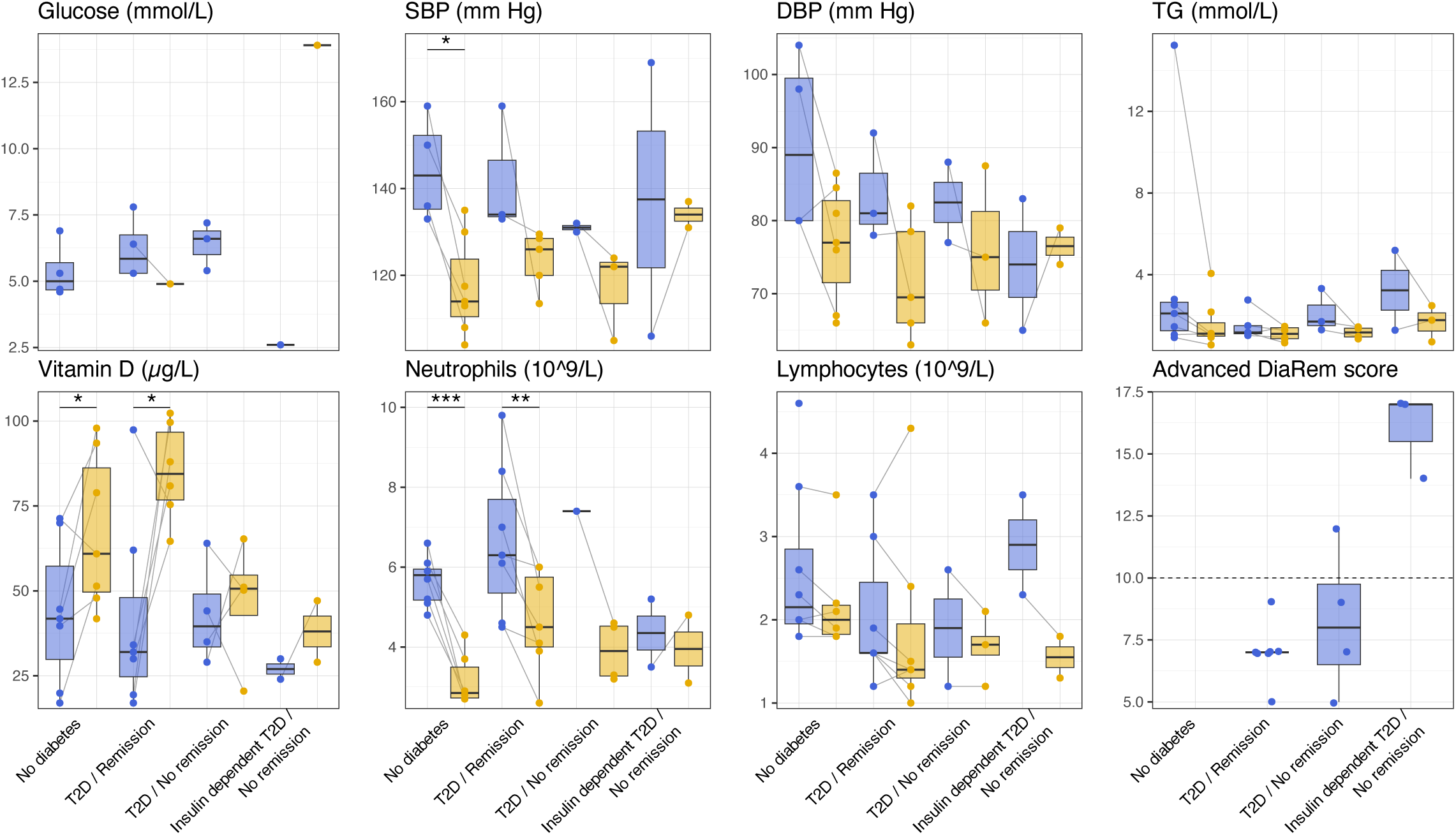
Boxplots illustrating the distribution of additional anthropometric and biochemical measurements across patient subgroups and visits. Legend similar to Figure 1B.

**Figure S2.**
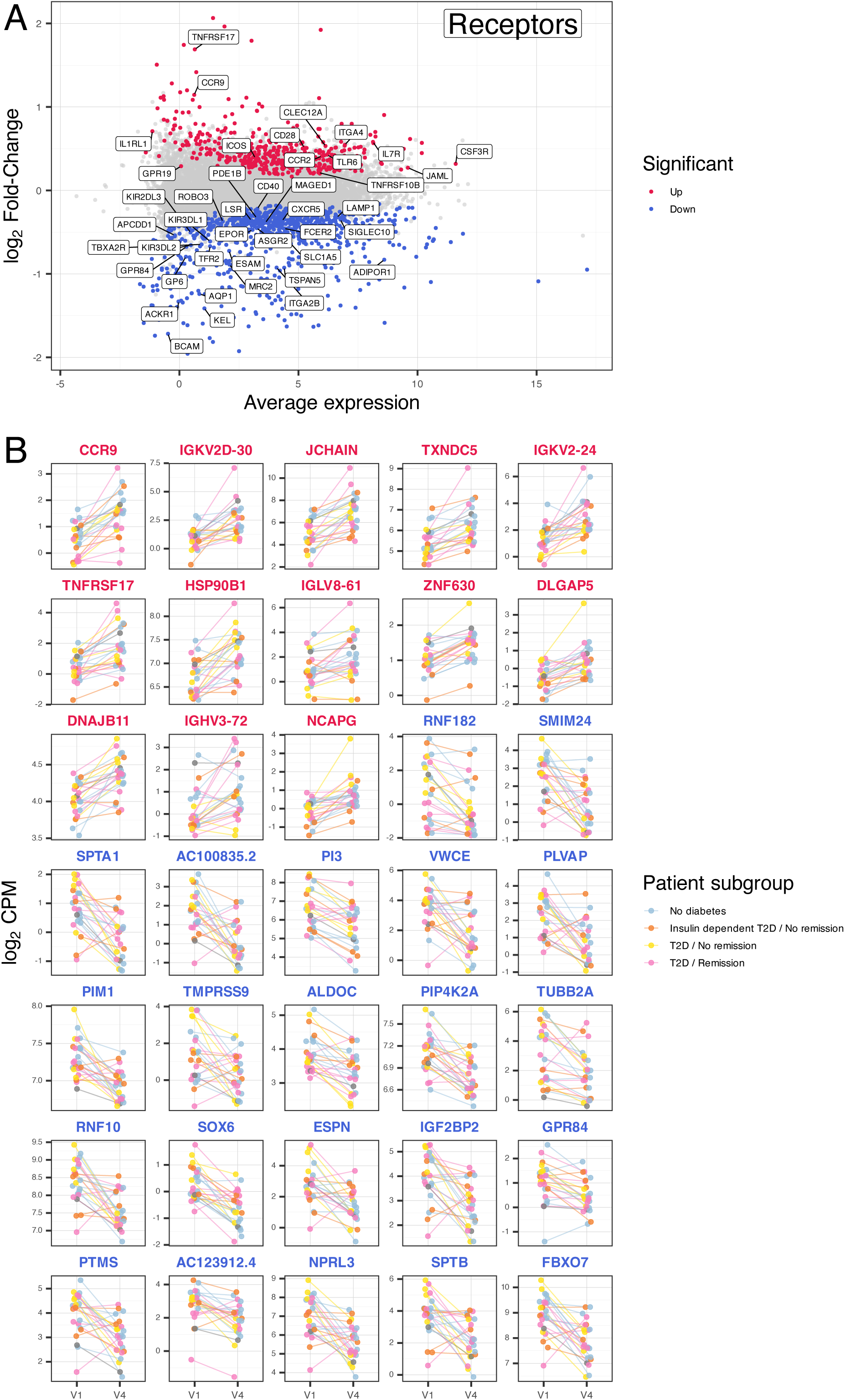
(A) Similar to Figure 3. Genes labelled with their symbol are coding for annotated receptors of ligands. (B) Normalized expression levels at visits 1 and 4 for the top differentially expressed genes (FDR 1%). Lines connect samples originating from the same patient. Dots and lines colour represent the patient subgroup. Two genes (BLOC1S5-TXNDC5 and IGKV2-30) were not displayed because they have high sequence similarity to other genes in the list (resp. TXNDC5 and IGKV2D-30) and were thus found differentially expressed based on the same sequencing reads.

**Figure S3.**
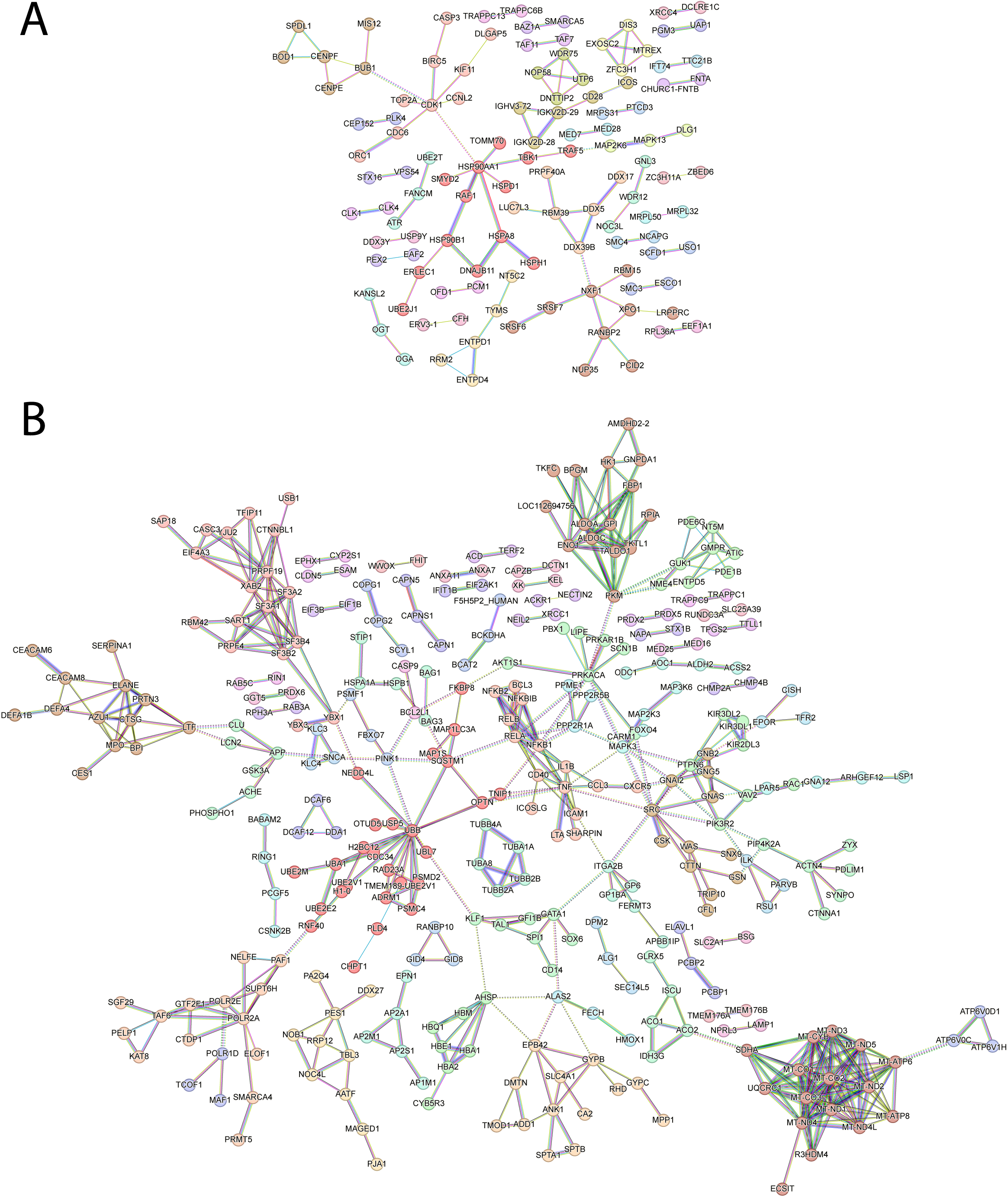
StringDB function association networks, showing high-confidence known and predicted interactions between proteins encoded by genes significantly up-regulated (A) or down-regulated (B) in the pre- vs. post-surgery differential expression analysis. Network nodes are proteins, coloured by their cluster membership. Network edges represent the predicted functional associations, and their colour represents the source of evidence for the associations (from curated databases in turquoise; experimentally determined in purple; from gene neighbourhood in green; from gene fusion in red; from gene co-occurrence in blue; from text mining in light green; and from protein homology in light blue). Associations within a cluster of proteins are depicted with plain lines, whereas associations across clusters are depicted with dotted lines.

**Figure S4.**
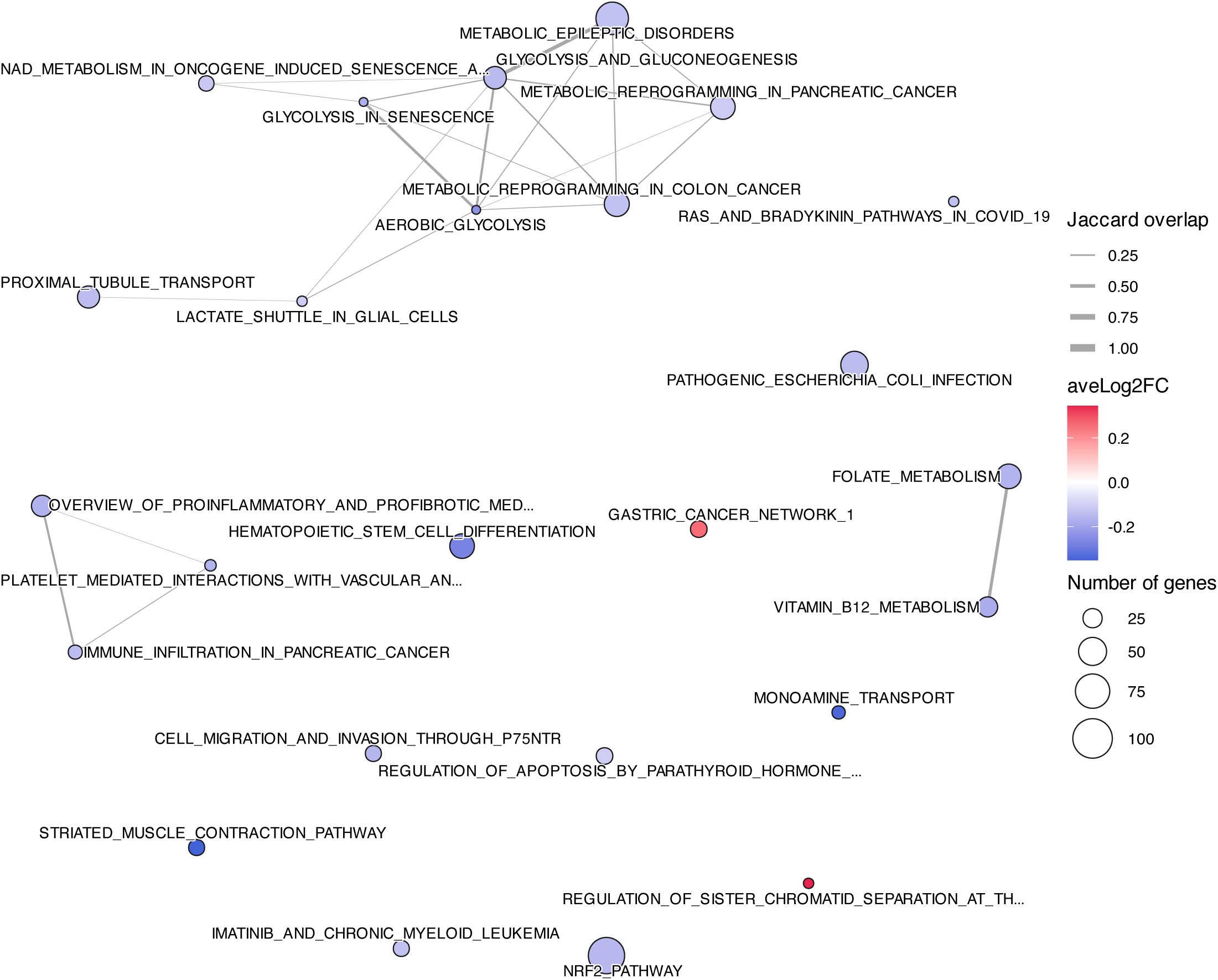
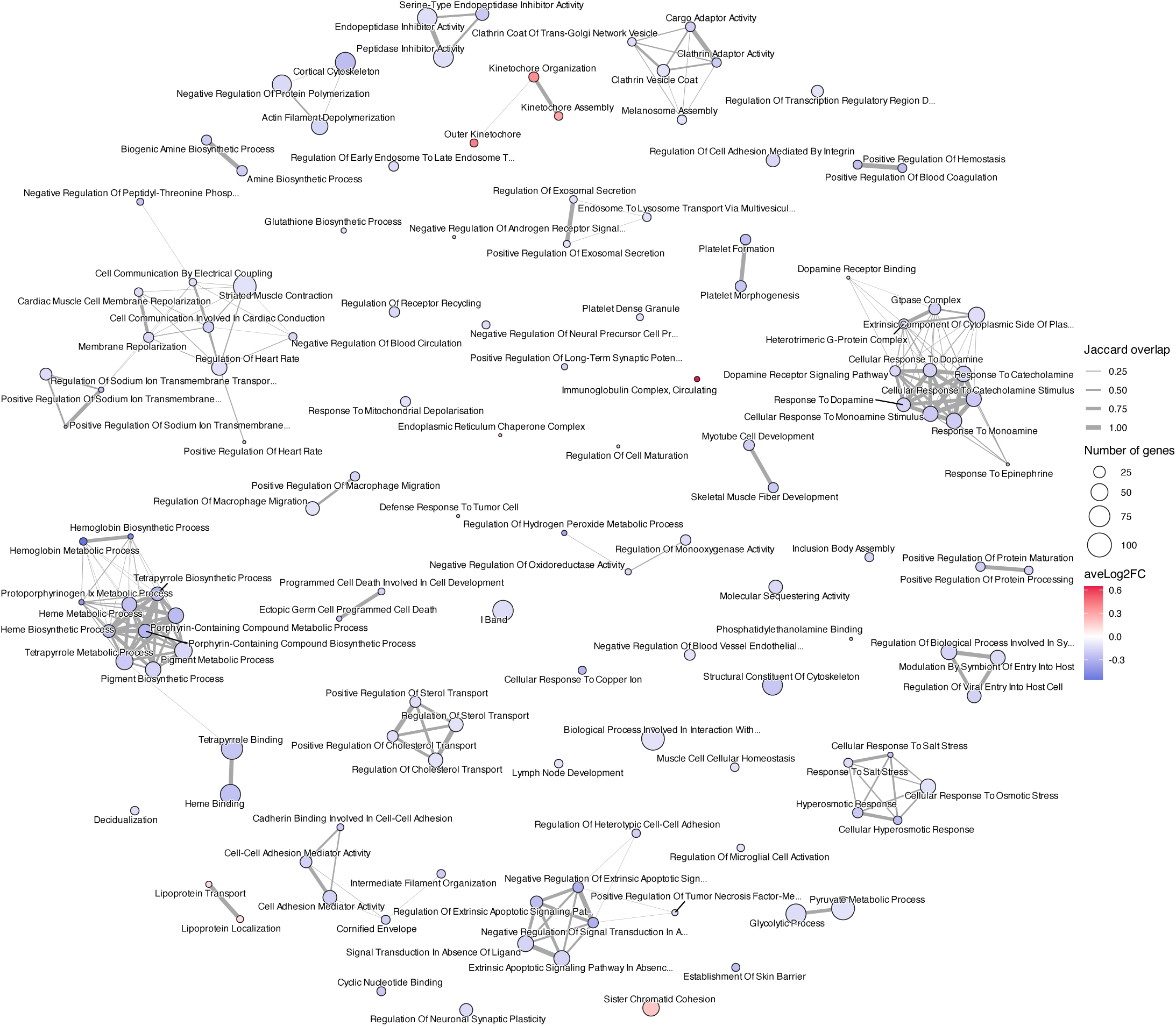
Enrichment maps similar to Figure 4, A) on the Wikipathways subset of canonical pathways from the MSigDB curated gene sets (C2) collection (full results shown in Table S4); B) on the Gene Ontology functional categories (full results shown in Table S5).

## Data Availability

Pre-processed RNA-seq data are available from GEO, under accession GSE273902. Raw data are not available due to patient privacy concerns. 
The code allowing reproducibility of the analyses presented in this paper is available
at https://github.com/julien-roux/Roux_et_al_bariatric_surgery_paper

https://www.ncbi.nlm.nih.gov/geo/query/acc.cgi?acc=GSE273902

https://github.com/julien-roux/Roux_et_al_bariatric_surgery_paper/tree/main

## Code availability

The code allowing reproducibility of the analyses presented in this paper is available at https://github.com/julien-roux/Roux_et_al_bariatric_surgery_paper.

## Acknowledgments

Calculations were performed at sciCORE (http://scicore.unibas.ch/) scientific computing centre at the University of Basel. We thank all patients for their contribution to the study. We thank Robert Ivanek, Anastasiya Börsch and Athimed El Taher from the DBM bioinformatics core facility at the University of Basel for useful discussions and feedback on the manuscript. We thank the authors of published transcriptome studies who shared their data. DS thanks Lukas Jeker for funding. Funding for sample collection was from the Imperial College BRC, and for RNA-sequencing was provided by the MRC. JB was funded by the Wellcome Trust. MRJ acknowledges funding from longITools, grant number 874739, 2020-2025. CCW and MRJ received funding from the EFSD Future Leaders Mentorship Programme for Clinical Diabetologists (educational grant from AstraZeneca). CCW acknowledges funding from the Swiss National Science Foundation (grant number 32003B_204937 / 2) and the Filling the Gap 2022-2023 – Siegenthaler Fellowship.

## Authors contributions

Conceptualization: JR, KHH, MRJ, AIB, CCW

Sample processing: JLB, SA

Methodology, formal analysis, visualization: JR, DS

Patient metadata curation: CCW, GH, DH, JR, DS

Writing: JR, with input from JB, DH, GH, KHH, MRJ, AIB and CCW

Funding acquisition: AIB, CCW

## Supplementary Text, Roux et al. 2025

### Description of the top differentially expressed genes

Among the most significant up-regulated genes (**Figure S2B**), numerous genes were specific to B cells (immunoglobulin variable genes from heavy and light chains, *JCHAIN, TNFRSF17, CCR9,* a receptor for CCL25/TECK expressed at the surface of gut-homing B and T cells [1]). CCR9 was previously documented in the context of obesity-induced diabetes [2]. In mice, its expression was shown to be reduced upon high-fat diet, and restored after diet-induced weight loss [3]. Other genes reflected potentially dysregulated cellular programs and pathways, such as an increase in heat shock response and genes related to protein folding (e.g., *DNAJB11, HSP90B1, TXNDC5*).

Genes involved in the regulation of the cell cycle were affected in both directions: *DLGAP5* and *NCAPG* were up-regulated, while *RNF10, RNF182, FBXO7* were down-regulated. Potentially also linked to cell cycle progression, *NPRL3,* a component of the GATOR1 complex which functions as a negative regulator of the mTORC1 pathway [4], was down-regulated, along with *PIM1,* known to have a positive regulator role on this complex by mediating inhibition of its component DEPDC5 [5].

Genes related to metabolism and its regulation were among the top down-regulated genes (**Figure S2B**)(e.g., *GPR84, TMPRSS9, PIP4K2A, ALDOC*). Some of these could reflect changes on the insulin metabolism pathway, such as *IGF2BP2*, an RNA-binding factor known to bind to the 5’-UTR of the insulin-like growth factor 2 mRNA, as well as *PIP4K2A*, known to negatively regulate insulin signaling [6, 7].

In the context of obesity or weight loss, ALDOC was shown to be down-regulated after weight loss [8]. SOX6 is a transcription factor known to regulate adipogenesis [9, 10]. PI3 encodes the anti-inflammatory protein Elafin, which protects the intestinal barrier function by inhibiting elastase activity.

Finally, we noted a decrease in expression of multiple cytoskeleton genes (e.g., ESPN, SPTA1, SPTB, TUBB2A).

### Functional network of genes differentially expressed after MBS

To explore the differential expression results, we used direct protein-protein interaction information gathered from the StringDB database [11] to create a network of functional associations of proteins produced by the up- and down-regulated genes. For up-regulated genes, the largest cluster of interacting proteins was made of proteins displaying chaperone activity (multiple HSP proteins, DNAJB1, ERLEC1) (**Figure S3A**). Another large cluster of proteins was associated with the regulation of mitosis and cell cycle (CDK1, TOP2A, DLGAP5, BIRC5, CDC6, KIF11, ORC1; BUB1, MIS12, SPDL1, BOD1, CENPE, CENPF; ESCO1, SMC3; NCAPG, SMC4). Other clusters were related to DNA repair (REV1, UBE2T, ATR), transcription initiation (TAF7, TAF11, components of the mediator complex MED7 and MED28), mRNA splicing (PRPF40A, RBM39, DDX5, DDX17, DDX39B, LUC7L3; CLK1, CLK4), RNA export from the nucleus (NXF1, XPO1, LRPPRC, NUP35, PCID2, RANBP2, SRSF7), ribosome biogenesis (ribosomal proteins, mitochondrial ribosomal proteins, protein associated with nucleolar RNAs such as NOP58 or UTP6, and components of the RNA exosome complex required for the maturation of these RNA species) and protein transport (SCFD1, USO1; STX16, VPS54). Finally, some clusters of proteins were related to lymphocyte cells (immunoglobulin genes and the proteins DCLRE1C and XRCC4 required for V(D)J recombination, the co-receptors CD28 and ICOS expressed at the surface of T cells and involved in interactions with B cells) and the regulation of inflammatory response (MAP2K6, MAPK13; TBK1, TRAF5; DDX3Y).

The network of interacting proteins produced by down-regulated genes featured clusters of proteins involved in immune regulation (**Figure S3B**). In particular, a large cluster of proteins included the subunits of the NF-κB complex (both canonical NFKB1 and RELA, and non-canonical NFKB2 and RELB [12]), some of their upstream regulators (NFKBIB, SHARPIN, the activating receptors LTBR, CD40 and ligands TNFα, LTA and IL1β) and downstream targets (BCL3, CCL3 [13], ICOSLG [14], CXCR5 [15], ICAM1). The canonical and non-canonical NF-κB pathways are tightly regulated by ubiquitination [16], and indeed a large cluster of proteins was centered around Ubiquitin (UBB) and enzymes related to ubiquitination, components of the 26S proteasome (ADRM1, PSMC4, PSMD2) and proteins known to be involved in NF-κB regulation (TNIP1, SQSTM1).

A large cluster of proteins was composed of proteins from the mitochondrial genome (MT-ATP6, MT-ATP8, MT-CO1, MT-CO2, MT-CO3, MT-CYB, MT-ND1, MT-ND2 MT-ND3, MT-ND4, MT-ND4L, MT-ND5) and from other proteins with mitochondrial function (mitochondrial SDHA, ECSIT; NME4, NT5M; ACO2, GLRX5, IDH3G, ISCU, UQCRC1).

Other clusters were related to the energy metabolism of cells, with proteins involved in glycolysis and gluconeogenesis (ALDOA, ALDOC, ENO1, FBP1, GNPDA1, GPI, HK1, LOC112694756, PKM, PIA, TALDO1, TKFC, TKTL1) or involved in the regulation of insulin signaling (GSK3A, PIP4K2A, FOXO4).

Additional clusters gathered proteins related to the regulation of transcription initiation and elongation (POLR2A, POLR2E, CTDP1, ELOF1, GTF2F1, KAT8, NELFE, PAF1, SMARCA4, SUPT6H, TAF6; POLR1D, MAF1, TCOF1), regulation of mRNA splicing (including multiple components of the spliceosome such as CASC3, CTNNBL1, EIF4A3, PRPF19, PRPF4, SART1, SF3A1, SF3A2, SF3B2, SF3B4, TFIP11, XAB2, YJU2, and RNA-binding proteins such as RBM42, YBX1, YBX3), regulation of ribosomal rRNA maturation (DDX27, NOB1, NOC4L, PA2G4, PES1, RRP12, TBL3) and protein chaperones (BAG1, BAG3, HSPA1A, HSPB1, STIP1).

A cluster of proteins was related to neutrophil activity (AZU1, CEACAM6, CEACAM8, DEFA1B, DEFA4, ELANE, PRTN3, LTF; LCN2), and multiple clusters were related to erythrocytes, including the haemoglobin subunits (HBA1, HBA2, HBE1, HBQ1, HBM), multiple proteins involved in erythrocyte differentiation and function (AHSP, CYB5R3; GATA1, KLF1, TAL1, ALAS2, FECH, HMOX1, GFI1B, SOX6), multiple erythrocyte membrane proteins (RHD, KEL, XK, GYPC, MPP1, ACKR) with components of the ankyrin-1 complex (ANK1, GYPB, EPB42, SLC4A1) linking the membrane to the spectrin-actin cytoskeletal network (SPTA1, SPTB, ADD1, DMTN) and thus a key element of the erythrocyte membrane stability and shape. Clusters of cytoskeletal proteins such as actin-associated proteins (e.g., ACTN4, CTNNA1, SYNPO, ZYX; CAPZB, DCTN1, TMOD1) and tubulins (TUBA1A, TUBA8, TUBB2A, TUBB2B, TUBB4A) could potentially be related to this erythrocyte pattern.

### Gene set enrichment analysis on Wikipathways pathways and Gene Ontology functional categories

Similar to pathway enrichment analysis using the Reactome database (**Figure 4)**, we used the Wikipathways database [17] to find significantly enriched pathways among genes differentially expressed after MBS (**Figure S4A; Table S4**). These included a down-regulation of “glycolysis and gluconeogenesis” (WP534), immune activation pathways (“immune infiltration in pancreatic cancer”, WP5285) and NF-κB signaling-related pathways such as vitamin B12 (WP1533) and folate metabolism (WP176). Two up-regulated pathways (“gastric cancer network 1”, WP2361 and “regulation of sister chromatid separation at the metaphase-anaphase transition”, WP4240) could suggest an increased cell proliferation.

The significantly enriched functional categories obtained when using the Gene Ontology database [18] are shown in **Figure S4B** (**Table S5**). Categories related to GPCR signal transduction were down-regulated. Additionally, a down-regulation of categories related to the cellular response to osmotic stress was observed. Categories related to B cells were up-regulated, while categories related to platelet formation were down-regulated. Some down-regulated functional categories related to the regulation of heart rate and to the regulation of cholesterol transport, while categories related to lipoprotein transport were up-regulated.

## References

1. Donath, M.Y. and S.E. Shoelson, Type 2 diabetes as an inflammatory disease. Nat Rev Immunol, 2011. 11(2): p. 98–107.

2. Hildebrandt, X., M. Ibrahim, and N. Peltzer, Cell death and inflammation during obesity: “Know my methods, WAT(son)”. Cell Death Differ, 2023. 30(2): p. 279–292.

3. van der Pouw Kraan, T.C., et al., Metabolic changes in type 2 diabetes are reflected in peripheral blood cells, revealing aberrant cytotoxicity, a viral signature, and hypoxia inducible factor activity. BMC Med Genomics, 2015. 8: p. 20.

4. Mahlangu, T., et al., A systematic review on the functional role of Th1/Th2 cytokines in type 2 diabetes and related metabolic complications. Cytokine, 2020. 126: p. 154892.

5. Govindasamy, C., A Review Emphasis on Imbalance of Th1/Th2 Cytokines in The Progression of Diabetes to Diabetic Related Complications. Oriental Journal Of Chemistry, 2024. 40(2): p. 355–361.

6. Zhang, Q., et al., VEGF levels in plasma in relation to metabolic control, inflammation, and microvascular complications in type-2 diabetes: A cohort study. Medicine, 2018. 97(15).

7. Cheng, L., et al., The effect of short-term intensive insulin therapy on circulating T cell subpopulations in patients with newly diagnosed type 2 diabetes mellitus. Diabetes Res Clin Pract, 2019. 149: p. 107–114.

8. Zhou, T., et al., Role of Adaptive and Innate Immunity in Type 2 Diabetes Mellitus. J Diabetes Res, 2018. 2018: p. 7457269.

9. SantaCruz-Calvo, S., et al., Adaptive immune cells shape obesity-associated type 2 diabetes mellitus and less prominent comorbidities. Nat Rev Endocrinol, 2022. 18(1): p. 23–42.

10. Tonyan, Z.N., et al., Overview of Transcriptomic Research on Type 2 Diabetes: Challenges and Perspectives. Genes (Basel), 2022. 13(7).

11. Choban, P.S., et al., Bariatric surgery for morbid obesity: Why, who, when, how, where, and then what? Cleveland Clinic Journal of Medicine, 2002. 69(11): p. 897.

12. Villarreal-Calderon, J.R., et al., Interplay between the Adaptive Immune System and Insulin Resistance in Weight Loss Induced by Bariatric Surgery. Oxid Med Cell Longev, 2019. 2019: p. 3940739.

13. Mingrone, G., et al., Bariatric-metabolic surgery versus conventional medical treatment in obese patients with type 2 diabetes: 5 year follow-up of an open-label, single-centre, randomised controlled trial. Lancet, 2015. 386(9997): p. 964–73.

14. Cosentino, C., et al., Efficacy and effects of bariatric surgery in the treatment of obesity: Network meta-analysis of randomized controlled trials. Nutr Metab Cardiovasc Dis, 2021. 31(10): p. 2815–2824.

15. Hage, K., et al., Type 2 diabetes remission after Roux-en-Y gastric bypass: a multicentered experience with long-term follow-up. Surg Obes Relat Dis, 2023. 19(12): p. 1339–1345.

16. Madsen, L.R., et al., Effect of Roux-en-Y gastric bypass surgery on diabetes remission and complications in individuals with type 2 diabetes: a Danish population-based matched cohort study. Diabetologia, 2019. 62(4): p. 611–620.

17. Villarreal-Calderon, J.R., et al., Reduced Th1 response is associated with lower glycolytic activity in activated peripheral blood mononuclear cells after metabolic and bariatric surgery. J Endocrinol Invest, 2021. 44(12): p. 2819–2830.

18. Askarpour, M., et al., Effect of Bariatric Surgery on Serum Inflammatory Factors of Obese Patients: a Systematic Review and Meta-Analysis. Obes Surg, 2019. 29(8): p. 2631–2647.

19. Villarreal-Calderon, J.R., et al., Metabolic shift precedes the resolution of inflammation in a cohort of patients undergoing bariatric and metabolic surgery. Sci Rep, 2021. 11(1): p. 12127.

20. Smith, E.P., et al., Altered glucose metabolism after bariatric surgery: What’s GLP-1 got to do with it? Metabolism, 2018. 83: p. 159–166.

21. Affinati, A.H., et al., Bariatric Surgery in the Treatment of Type 2 Diabetes. Curr Diab Rep, 2019. 19(12): p. 156.

22. Kratz, A., et al., Case records of the Massachusetts General Hospital. Weekly clinicopathological exercises. Laboratory reference values. N Engl J Med, 2004. 351(15): p. 1548–63.

23. Stevens, J.R., et al., Power in pairs: assessing the statistical value of paired samples in tests for differential expression. BMC Genomics, 2018. 19(1): p. 953.

24. Gomes, I., et al., Identification of GPR83 as the receptor for the neuroendocrine peptide PEN. Science Signaling, 2016. 9(425): p. ra43-ra43.

25. Roh, J., et al., Intermedin is a calcitonin/calcitonin gene-related peptide family peptide acting through the calcitonin receptor-like receptor/receptor activity-modifying protein receptor complexes. J Biol Chem, 2004. 279(8): p. 7264–74.

26. Kazak, L., et al., Ablation of adipocyte creatine transport impairs thermogenesis and causes diet-induced obesity. Nat Metab, 2019. 1(3): p. 360–370.

27. Peleli, M., et al., Dietary nitrate attenuates high-fat diet-induced obesity via mechanisms involving higher adipocyte respiration and alterations in inflammatory status. Redox Biol, 2020. 28: p. 101387.

28. Yue, J.T., et al., Inhibition of glycine transporter-1 in the dorsal vagal complex improves metabolic homeostasis in diabetes and obesity. Nat Commun, 2016. 7: p. 13501.

29. Benedet, P.O., et al., CD248 promotes insulin resistance by binding to the insulin receptor and dampening its insulin-induced autophosphorylation. EBioMedicine, 2024. 99: p. 104906.

30. Kalis, M., et al., alpha 1-antitrypsin enhances insulin secretion and prevents cytokine-mediated apoptosis in pancreatic beta-cells. Islets, 2010. 2(3): p. 185–9.

31. Ragolia, L., C.E. Hall, and T. Palaia, Lipocalin-type prostaglandin D(2) synthase stimulates glucose transport via enhanced GLUT4 translocation. Prostaglandins Other Lipid Mediat, 2008. 87(1-4): p. 34–41.

32. Ragolia, L., et al., Accelerated glucose intolerance, nephropathy, and atherosclerosis in prostaglandin D2 synthase knock-out mice. J Biol Chem, 2005. 280(33): p. 29946–55.

33. Wang, Y., et al., Clusterin is closely associated with adipose tissue insulin resistance. Diabetes Metab Res Rev, 2023. 39(7): p. e3688.

34. Yadav, N. and S.-Y. Kim, Transglutaminase2: An Enduring Enzyme in Diabetes and Age-Related Metabolic Diseases. Kinases and Phosphatases, 2024. 2(1): p. 67–91.

35. Narvekar, P., et al., Liver-specific loss of lipolysis-stimulated lipoprotein receptor triggers systemic hyperlipidemia in mice. Diabetes, 2009. 58(5): p. 1040–9.

36. Norouzirad, R., P. Gonzalez-Muniesa, and A. Ghasemi, Hypoxia in Obesity and Diabetes: Potential Therapeutic Effects of Hyperoxia and Nitrate. Oxid Med Cell Longev, 2017. 2017: p. 5350267.

37. Reimold, A.M., et al., Plasma cell differentiation requires the transcription factor XBP-1. Nature, 2001. 412(6844): p. 300–307.

38. Piperi, C., C. Adamopoulos, and A.G. Papavassiliou, XBP1: A Pivotal Transcriptional Regulator of Glucose and Lipid Metabolism. Trends Endocrinol Metab, 2016. 27(3): p. 119–122.

39. Szklarczyk, D., et al., The STRING database in 2023: protein-protein association networks and functional enrichment analyses for any sequenced genome of interest. Nucleic Acids Res, 2023. 51(D1): p. D638–D646.

40. Milacic, M., et al., The Reactome Pathway Knowledgebase 2024. Nucleic Acids Res, 2024. 52(D1): p. D672–D678.

41. Vallese, F., et al., Architecture of the human erythrocyte ankyrin-1 complex. Nat Struct Mol Biol, 2022. 29(7): p. 706–718.

42. Agrawal, A., et al., WikiPathways 2024: next generation pathway database. Nucleic Acids Res, 2024. 52(D1): p. D679–D689.

43. Gene Ontology Consortium, et al., The Gene Ontology knowledgebase in 2023. Genetics, 2023. 224(1).

44. Aran, D., Z. Hu, and A.J. Butte, xCell: digitally portraying the tissue cellular heterogeneity landscape. Genome Biol, 2017. 18(1): p. 220.

45. Homuth, G., et al., Extensive alterations of the whole-blood transcriptome are associated with body mass index: results of an mRNA profiling study involving two large population-based cohorts. BMC Med Genomics, 2015. 8: p. 65.

46. Kalafati, M., et al., An interferon-related signature characterizes the whole blood transcriptome profile of insulin-resistant individuals—the CODAM study. Genes & Nutrition, 2021. 16(1): p. 22.

47. Wesolowska-Andersen, A., et al., Four groups of type 2 diabetes contribute to the etiological and clinical heterogeneity in newly diagnosed individuals: An IMI DIRECT study. Cell Rep Med, 2022. 3(1): p. 100477.

48. de Klerk, J.A., et al., Altered blood gene expression in the obesity-related type 2 diabetes cluster may be causally involved in lipid metabolism: a Mendelian randomisation study. Diabetologia, 2023. 66(6): p. 1057–1070.

49. Liu, N., et al., Investigating the change in gene expression profile of blood mononuclear cells post-laparoscopic sleeve gastrectomy in Chinese obese patients. Front Endocrinol (Lausanne), 2023. 14: p. 1049484.

50. Rashid, M., et al., Transcriptome Changes and Metabolic Outcomes After Bariatric Surgery in Adults With Obesity and Type 2 Diabetes. J Endocr Soc, 2023. 8(1): p. bvad159.

51. Berisha, S.Z., et al., Changes in whole blood gene expression in obese subjects with type 2 diabetes following bariatric surgery: a pilot study. PLoS One, 2011. 6(3): p. e16729.

52. Sandoval, D.A. and M.E. Patti, Glucose metabolism after bariatric surgery: implications for T2DM remission and hypoglycaemia. Nat Rev Endocrinol, 2023. 19(3): p. 164–176.

53. Rohm, T.V., et al., Inflammation in obesity, diabetes, and related disorders. Immunity, 2022. 55(1): p. 31–55.

54. Schmidt, V., et al., Obesity-Mediated Immune Modulation: One Step Forward, (Th)2 Steps Back. Front Immunol, 2022. 13: p. 932893.

55. Luck, H., et al., Gut-associated IgA(+) immune cells regulate obesity-related insulin resistance. Nat Commun, 2019. 10(1): p. 3650.

56. Lautenbach, A., et al., Long-Term Improvement of Chronic Low-Grade Inflammation After Bariatric Surgery. Obes Surg, 2021. 31(7): p. 2913–2920.

57. Barbosa, P., et al., Bariatric Surgery Induces Alterations in the Immune Profile of Peripheral Blood T Cells. Biomolecules, 2024. 14(2).

58. Zhang, P., K. Watari, and M. Karin, Innate immune cells link dietary cues to normal and abnormal metabolic regulation. Nat Immunol, 2025. 26(1): p. 29–41.

59. Nijhuis, J., et al., Neutrophil activation in morbid obesity, chronic activation of acute inflammation. Obesity (Silver Spring), 2009. 17(11): p. 2014–8.

60. Kleinstein, S.E., et al., Transcriptomics of type 2 diabetic and healthy human neutrophils. BMC Immunol, 2021. 22(1): p. 37.

61. Lin, Q., et al., Abnormal Peripheral Neutrophil Transcriptome in Newly Diagnosed Type 2 Diabetes Patients. J Diabetes Res, 2020. 2020: p. 9519072.

62. Lo, T., et al., Early Changes in Immune Cell Count, Metabolism, and Function Following Sleeve Gastrectomy: A Prospective Human Study. J Clin Endocrinol Metab, 2022. 107(2): p. e619–e630.

63. Poitou, C., et al., Bariatric Surgery Induces Disruption in Inflammatory Signaling Pathways Mediated by Immune Cells in Adipose Tissue: A RNA-Seq Study. PLoS One, 2015. 10(5): p. e0125718.

64. Cummings, D.E. and M.H. Shannon, Roles for Ghrelin in the Regulation of Appetite and Body Weight. Archives of Surgery, 2003. 138(4): p. 389–396.

65. Pournaras, D.J. and C.W. le Roux, Ghrelin and metabolic surgery. Int J Pept, 2010. 2010.

66. Wijngaarden, L.H., et al., T and B Cell Composition and Cytokine Producing Capacity Before and After Bariatric Surgery. Front Immunol, 2022. 13: p. 888278.

67. Sachan, A., et al., An immediate post op and follow up assessment of circulating adipo-cytokines after bariatric surgery in morbid obesity. Metabol Open, 2022. 13: p. 100147.

68. Chen, M., et al., Hematological disorders following gastric bypass surgery: emerging concepts of the interplay between nutritional deficiency and inflammation. Biomed Res Int, 2013. 2013: p. 205467.

69. McShane, E., et al., A kinetic dichotomy between mitochondrial and nuclear gene expression processes. Mol Cell, 2024. 84(8): p. 1541–1555 e11.

70. Papadakis, G.E., et al., Multiomics unravels the complexity of male obesity: a prospective observational study. J Transl Med, 2025. 23(1): p. 138.

71. Voros, C., et al., Nitrate-Nitrite-Nitric Oxide Pathway, Oxidative Stress, and Fertility Outcomes in Morbidly Obese Women Following Bariatric Surgery: A Systematic Review. Biomedicines, 2024. 13(1).

72. Christakoudi, S., et al., Associations of obesity and body shape with erythrocyte and reticulocyte parameters in the UK Biobank cohort. BMC Endocr Disord, 2023. 23(1): p. 161.

73. Fleiss, J.L., Design and Analysis of Clinical Experiments. 2011: Wiley.

74. Riddle, M.C., et al., Consensus Report: Definition and Interpretation of Remission in Type 2 Diabetes. Diabetes Care, 2021. 44(10): p. 2438–44.

75. Aron-Wisnewsky, J., et al., The advanced-DiaRem score improves prediction of diabetes remission 1 year post-Roux-en-Y gastric bypass. Diabetologia, 2017. 60(10): p. 1892–1902.

76. Dobin, A., et al., STAR: ultrafast universal RNA-seq aligner. Bioinformatics, 2013. 29(1): p. 15–21.

77. Liao, Y., G.K. Smyth, and W. Shi, featureCounts: an efficient general purpose program for assigning sequence reads to genomic features. Bioinformatics, 2014. 30(7): p. 923–30.

78. Huber, W., et al., Orchestrating high-throughput genomic analysis with Bioconductor. Nat Meth, 2015. 12(2): p. 115–121.

79. Ballman, K.V., et al., Faster cyclic loess: normalizing RNA arrays via linear models. Bioinformatics, 2004. 20(16): p. 2778–86.

80. Law, C., et al., voom: precision weights unlock linear model analysis tools for RNA-seq read counts. Genome Biology, 2014. 15(2): p. R29.

81. Wu, D., et al., ROAST: rotation gene set tests for complex microarray experiments. Bioinformatics, 2010. 26(17): p. 2176–82.

82. Liberzon, A., et al., The Molecular Signatures Database (MSigDB) hallmark gene set collection. Cell Syst, 2015. 1(6): p. 417–425.

83. Jin, S., et al., Inference and analysis of cell-cell communication using CellChat. Nat Commun, 2021. 12(1): p. 1088.

84. Shao, X., et al., CellTalkDB: a manually curated database of ligand-receptor interactions in humans and mice. Brief Bioinform, 2021. 22(4).

85. Troulé, K., et al., CellPhoneDB v5: inferring cell-cell communication from single-cell multiomics data. arXiv e-prints, 2023: p. arXiv:2311.04567.

86. Badia, I.M.P., et al., decoupleR: ensemble of computational methods to infer biological activities from omics data. Bioinform Adv, 2022. 2(1): p. vbac016.

87. Turei, D., T. Korcsmaros, and J. Saez-Rodriguez, OmniPath: guidelines and gateway for literature-curated signaling pathway resources. Nat Methods, 2016. 13(12): p. 966–967.

## References

1. Zabel, B.A., et al., Human G Protein–Coupled Receptor Gpr-9-6/Cc Chemokine Receptor 9 Is Selectively Expressed on Intestinal Homing T Lymphocytes, Mucosal Lymphocytes, and Thymocytes and Is Required for Thymus-Expressed Chemokine–Mediated Chemotaxis. Journal of Experimental Medicine, 1999. 190(9): p. 1241–1256.

2. Amiya, T., et al., C-C motif chemokine receptor 9 regulates obesity-induced insulin resistance via inflammation of the small intestine in mice. Diabetologia, 2021. 64(3): p. 603–617.

3. Park, C., et al., Obesity Modulates Intestinal Intraepithelial T Cell Persistence, CD103 and CCR9 Expression, and Outcome in Dextran Sulfate Sodium-Induced Colitis. J Immunol, 2019. 203(12): p. 3427–3435.

4. Shen, K., et al., Architecture of the human GATOR1 and GATOR1-Rag GTPases complexes. Nature, 2018. 556(7699): p. 64–69.

5. Padi, S.K.R., et al., Phosphorylation of DEPDC5, a component of the GATOR1 complex, releases inhibition of mTORC1 and promotes tumor growth. Proc Natl Acad Sci U S A, 2019. 116(41): p. 20505–20510.

6. Wang, D.G., et al., PIP4Ks Suppress Insulin Signaling through a Catalytic-Independent Mechanism. Cell Rep, 2019. 27(7): p. 1991–2001 e5.

7. Sharma, S., et al., Phosphatidylinositol 5 Phosphate 4-Kinase Regulates Plasma-Membrane PIP(3) Turnover and Insulin Signaling. Cell Rep, 2019. 27(7): p. 1979–1990 e7.

8. Verhoef, S.P., et al., Physiological response of adipocytes to weight loss and maintenance. PLoS One, 2013. 8(3): p. e58011.

9. Leow, S.C., et al., The transcription factor SOX6 contributes to the developmental origins of obesity by promoting adipogenesis. Development, 2016. 143(6): p. 950–61.

10. VanHook, A.M., SOX to be fat. Science Signaling, 2016. 9(420): p. ec66–ec66.

11. Szklarczyk, D., et al., The STRING database in 2023: protein-protein association networks and functional enrichment analyses for any sequenced genome of interest. Nucleic Acids Res, 2023. 51(D1): p. D638–D646.

12. Sun, S.C., Non-canonical NF-kappaB signaling pathway. Cell Res, 2011. 21(1): p. 71–85.

13. Sindhu, S., et al., MIP-1alpha Expression Induced by Co-Stimulation of Human Monocytic Cells with Palmitate and TNF-alpha Involves the TLR4-IRF3 Pathway and Is Amplified by Oxidative Stress. Cells, 2020. 9(8).

14. Hu, H., et al., Noncanonical NF-kappaB regulates inducible costimulator (ICOS) ligand expression and T follicular helper cell development. Proc Natl Acad Sci U S A, 2011. 108(31): p. 12827–32.

15. Wei, C., et al., CD40 Signaling Promotes CXCR5 Expression in B Cells via Noncanonical NF-kappaB Pathway Activation. J Immunol Res, 2020. 2020: p. 1859260.

16. Chen, Z.J., Ubiquitin signalling in the NF-κB pathway. Nature Cell Biology, 2005. 7(8): p. 758–765.

17. Agrawal, A., et al., WikiPathways 2024: next generation pathway database. Nucleic Acids Res, 2024. 52(D1): p. D679–D689.

18. Gene Ontology Consortium, et al., The Gene Ontology knowledgebase in 2023. Genetics, 2023. 224(1).

